# Development and Multinational Validation of an Ensemble Deep Learning Algorithm for Detecting and Predicting Structural Heart Disease Using Noisy Single-lead Electrocardiograms

**DOI:** 10.1101/2024.10.07.24314974

**Authors:** Arya Aminorroaya, Lovedeep S Dhingra, Aline Pedroso Camargos, Sumukh Vasisht Shankar, Andreas Coppi, Akshay Khunte, Murilo Foppa, Luisa CC Brant, Sandhi M Barreto, Antonio Luiz P Ribeiro, Harlan M Krumholz, Evangelos K Oikonomou, Rohan Khera

## Abstract

**Background and Aims:** AI-enhanced 12-lead ECG can detect a range of structural heart diseases (SHDs) but has a limited role in community-based screening. We developed and externally validated a noise-resilient single-lead AI-ECG algorithm that can detect SHD and predict the risk of their development using wearable/portable devices.

**Methods:** Using 266,740 ECGs from 99,205 patients with paired echocardiographic data at Yale New Haven Hospital, we developed ADAPT-HEART, a noise-resilient, deep-learning algorithm, to detect SHD using lead I ECG. SHD was defined as a composite of LVEF<40%, moderate or severe left-sided valvular disease, and severe LVH. ADAPT-HEART was validated in four community hospitals in the US, and the population-based cohort of ELSA-Brasil. We assessed the model’s performance as a predictive biomarker among those without baseline SHD across hospital-based sites and the UK Biobank.

**Results:** The development population had a median age of 66 [IQR, 54-77] years and included 49,947 (50.3%) women, with 18,896 (19.0%) having any SHD. ADAPT-HEART had an AUROC of 0.879 (95% CI, 0.870-0.888) with good calibration for detecting SHD in the test set, and consistent performance in hospital-based external sites (AUROC: 0.852-0.891) and ELSA-Brasil (AUROC: 0.859). Among those without baseline SHD, high vs. low ADAPT-HEART probability conferred a 2.8- to 5.7-fold increase in the risk of future SHD across data sources (all P<0.05).

**Conclusions:** We propose a novel model that detects and predicts a range of SHDs from noisy single-lead ECGs obtainable on portable/wearable devices, providing a scalable strategy for community-based screening and risk stratification for SHD.

## INTRODUCTION

Early diagnosis of structural heart disease (SHD) enables the timely initiation of therapies that can improve disease trajectory and patient outcomes.^1–6^ While a majority of SHDs are characterized by a long presymptomatic phase,^5–8^ there is currently no systematic approach for detecting SHD before symptom onset. This challenge arises from the lack of feasible screening strategies that can be deployed at scale without requiring advanced imaging or detailed healthcare evaluation.^9,10^ Currently, the diagnosis of SHD requires referral for an echocardiogram or other advanced cardiac imaging, with no specific guidance on which asymptomatic patients should be referred.^11^ The increasing availability of wearable and portable devices, many of which can capture single-lead ECGs, provides a low-cost cardiac diagnostic modality for a broad population.^12,13^ Nonetheless, there are no discernible signatures for detecting SHD from portable single-lead ECGs.

The advent of artificial intelligence (AI) algorithms has enabled the detection of subtle, human-unreadable digital signatures of distinct SHDs using 12-lead electrocardiograms (ECG).^14,15^ While such advances in AI enable a new role for ECGs as a modality for SHD screening, reliance on 12-lead ECGs obtained in clinical settings limits the scale and impact of such a screening strategy.^14,16^ Specifically, this strategy excludes individuals who have not undergone such testing, either due to the absence of clinical indications or lack of access. Conversely, portable and wearable devices enable the acquisition of single-lead ECGs from a broader population.^12,13^ We have previously demonstrated that left ventricular systolic dysfunction (LVSD), a key SHD, can be accurately identified on a noisy lead I ECG obtainable on wearable and portable devices.^17^ Nevertheless, detecting individual conditions in a community setting is challenging due to their low prevalence, with a high rate of false positives, even with high-performing models.^16^ This obstacle can be addressed by using single-lead ECGs to detect a composite of multiple SHDs, which would result in fewer false positives due to the higher overall prevalence of the composite SHD. This approach could represent an objective and scalable strategy for referral for cardiac imaging to detect SHD.

To fully leverage the promise of AI-ECG for transforming community-level ECG screening for SHD, we sought to develop and externally validate a noise-resilient deep learning algorithm to identify a range of clinically actionable SHDs spanning LVSD, moderate or severe left-sided valvular disease, and severe left ventricular hypertrophy (sLVH), using single-lead ECGs. We also explored the predictive role of this algorithm for the risk of incident SHD.

## METHODS

### Data Sources

We developed the model using ECG paired with echocardiographic data from the Yale New Haven Hospital (YNHH) during 2015-2023 (**Figure S1**). YNHH serves a large and diverse population in New Haven, one of the most representative counties in the US, and across Connecticut.^18^ We accessed 2.0 million ECG waveforms containing raw voltage data from all 12 leads of a standard clinical ECG. The echocardiographic data consisted of 335,184 transthoracic echocardiograms (TTE) with structured reports. For external validation, we obtained data from four geographically distinct community hospitals in the Yale New Haven Health System, covering other towns and regions in Southern Connecticut and Rhode Island. These include the Bridgeport Hospital, Greenwich Hospital, Lawrence + Memorial Hospital, and Westerly Hospital. To prevent any representation of the same patients across development and external validation datasets, we excluded patients from the external validation sites whose data had been used for model development. In addition to the hospital-based cohorts, we assessed the external validity of the model in the Brazilian Longitudinal Study of Adult Health (ELSA-Brasil), the largest prospective population-based cohort in Brazil with protocolized ECG and TTEs.^19^

To evaluate the predictive significance of the model, in addition to hospital-based cohorts, we included participants from the UK Biobank (UKB). UKB is the largest population-based cohort from the UK where participants underwent protocolized ECGs during 2014-2020 with linked data from the national death registry as well as the comprehensive electronic health records (EHR) data from the National Health Service England.^20^

### Study Population

To develop the model and assess its external validity, we included individuals from the clinical sites and ELSA-Brasil. From the clinical sites, we identified patients with at least one pair of ECG-TTE occurring within a 30-day window. Each ECG was paired with at most one TTE, while each TTE could have been paired with multiple ECGs. In cases where multiple TTEs were obtained within 30 days of an ECG, only the TTE closest to the ECG was included. Conversely, if multiple ECGs were recorded within 30 days of a TTE, we included up to five ECGs per patient in the development set to avoid over-representation of patients with frequent health encounters. We excluded information from patients who had a history of cardiac procedures at the time of ECG, including coronary artery bypass grafting, aortic or mitral valve procedures, left ventricular assist device implantation, heart transplant, alcohol septal ablation, and ventricular myectomy (**Figure S1**). From ELSA-Brasil, we included all participants who underwent a simultaneous ECG and TTE at their baseline visit during 2008-2010.

To assess the predictive role of the algorithm in predicting incident SHD, we included individuals from the clinical sites and the UKB. From the clinical sites, we identified patients with an ECG during 2013-2023 at YNHH and the four community hospitals. We excluded those with SHD based on TTE, heart failure based on diagnosis codes, or left-sided valve replacement or repair based on procedure codes before the index date (**Table S1**). In the clinical sites, diagnosis codes were recorded as the International Classification of Diseases, Tenth Revision, Clinical Modification (ICD-10-CM), and procedure codes were recorded as the International Classification of Diseases, Tenth Revision, Procedure Coding System (ICD-10-PCS) or Current Procedural Terminology 4 (CPT4). The index date was defined as the date of the first ECG acquisition at any time after one year from their first health encounter. This one-year blanking period was included to ensure prevalent SHD was not misclassified as incident SHD. We also excluded those from YNHH whose data were used for model development.

For predictive assessment in the UKB, we identified participants who received protocolized ECG during 2014-2021. We excluded those with a history of heart failure or left-sided valvular disease based on diagnosis codes, as well as those who had undergone left-sided valve replacement or repair based on procedure codes, before their ECG acquisition (**Table S1**). Diagnoses were coded in the International Classification of Diseases, Ninth and Tenth Revisions (ICD-9 and ICD-10), and procedures were coded in the Office of Population Censuses and Surveys Classification of Interventions and Procedures, versions 3 and 4 (OPCS-3 and OPCS-4). This information was obtained from the linkage of the UKB with the national EHR, predating the UKB enrollment in 2006, to ensure complete capture of baseline SHD status.

### Study Covariates

We defined SHD as the TTE-defined presence of a composite of LVSD, moderate or severe left-sided valvular disease, and/or sLVH. LVSD was defined as a left ventricular ejection fraction (LVEF) <40%. Moderate or severe left-sided valvular diseases were identified by the presence of any moderate or severe aortic regurgitation (AR), aortic stenosis (AS), mitral regurgitation (MR), or mitral stenosis (MS). We characterized sLVH by an interventricular septal diameter at end-diastole (IVSd) greater than 15 mm with concomitant moderate or severe left ventricular diastolic dysfunction (LVDD). All echocardiographic indices were measured according to the American Society of Echocardiography guidelines.^11^ IVSd and LVEF were measured objectively, while the severity of LVDD and valvular disease were graded by the reading cardiologist, based on the guidelines.^11^ LVEF was defined using three-dimensional echo, Simpson’s biplane, or visual estimation methods. The valvular component of SHD entailed moderate, moderate to severe, or severe stenotic or regurgitant disorders of aortic or mitral valves. We also evaluated an alternative definition for SHD that consists of LVSD and sLVH, but included severe, instead of moderate or severe, left-sided valvular disease.

### Development of an Ensemble Noise-adapted Model

We randomly split the included ECGs into training, internal validation, and held-out test sets with a ratio of 85:5:10 at the patient level to avoid train-test contamination. In the training set, we retained up to five random ECGs per patient to ensure the adequacy of training data, as noted above. However, the validation and test sets included only one random ECG per patient to avoid any artificial inflation or deflation of the model’s performance. Similarly, we included one random ECG per patient from the hospital-based external validation sites.

We employed a standard signal preprocessing strategy to extract the signal data from lead I - representing the standard lead captured by portable devices - of 12-lead ECG recordings in the YNHH development set. However, the lead I isolated from clinical 12-lead ECGs is often less noisy than real-world lead I ECGs acquired using portable and wearable devices.^21^ To account for these differences, we adopted our previously developed method to construct noise-resilient algorithms.^17^ Briefly, to conform clinical ECGs with portable ECGs, we augmented ECGs in the training set using random Gaussian noise, while we tested the model on clean ECGs that were not noised (**Supplemental Methods**). Algorithms trained using this approach have retained their performance when tested on lead I ECGs, even when augmented with real-world noise from portable devices.^17^

The model development involved a two-step approach where we first trained separate deep learning algorithms to detect individual SHD components, and then combined their outputs with the patient’s age and sex to detect the composite SHD label. To train label-specific algorithms, we employed a convolutional neural network (CNN) architecture with excellent discrimination for detecting LVSD from single-lead ECG.^17^ Leveraging this architecture and transfer learning, we trained six distinct CNN models to predict LVSD, moderate or severe left-sided valvular disease, moderate or severe AR, moderate or severe AS, moderate or severe MR, and sLVH (**Supplemental Methods**). All models were trained using all ECGs in the training set. However, given the low prevalence of sLVH (<1%), we adopted a case-control training strategy with age- and sex-matching for the sLVH model to ensure learning specific ECG signatures of sLVH. For the sLVH training set, each ECG from patients with sLVH was matched with 10 ECGs from patients without sLVH, ensuring that they had the same sex and were within a 5-year age window. To further improve the model’s ability to learn sLVH ECG signatures, we trained the sLVH model on extreme phenotypes, excluding intermediate ones. For training, positive cases were defined by the sLVH criteria (IVSd >15 mm and moderate or severe LVDD), and negative cases were defined by an IVSd <12 mm in the absence of moderate or severe LVDD. The model’s performance was then assessed based on the presence or absence of sLVH. Therefore, while the sLVH model was trained on extreme phenotypes, it was tested against the full spectrum of phenotypes in the internal validation and held-out test sets.

Leveraging an ensemble learning strategy, we developed ADAPT-HEART (AI Deep learning for Adapting Portable Technology in HEART disease detection) using extreme gradient boosting to predict the composite SHD based on the CNN models’ output probabilities, as well as the patient’s age and sex as predictive features (**Figure 1**). The ensemble model’s performance was evaluated in the held-out test set and external validation sets. For the predictive assessment, we deployed the ensemble algorithm to obtain the probability of concomitant SHD from corresponding ECGs.

**Figure 1.**
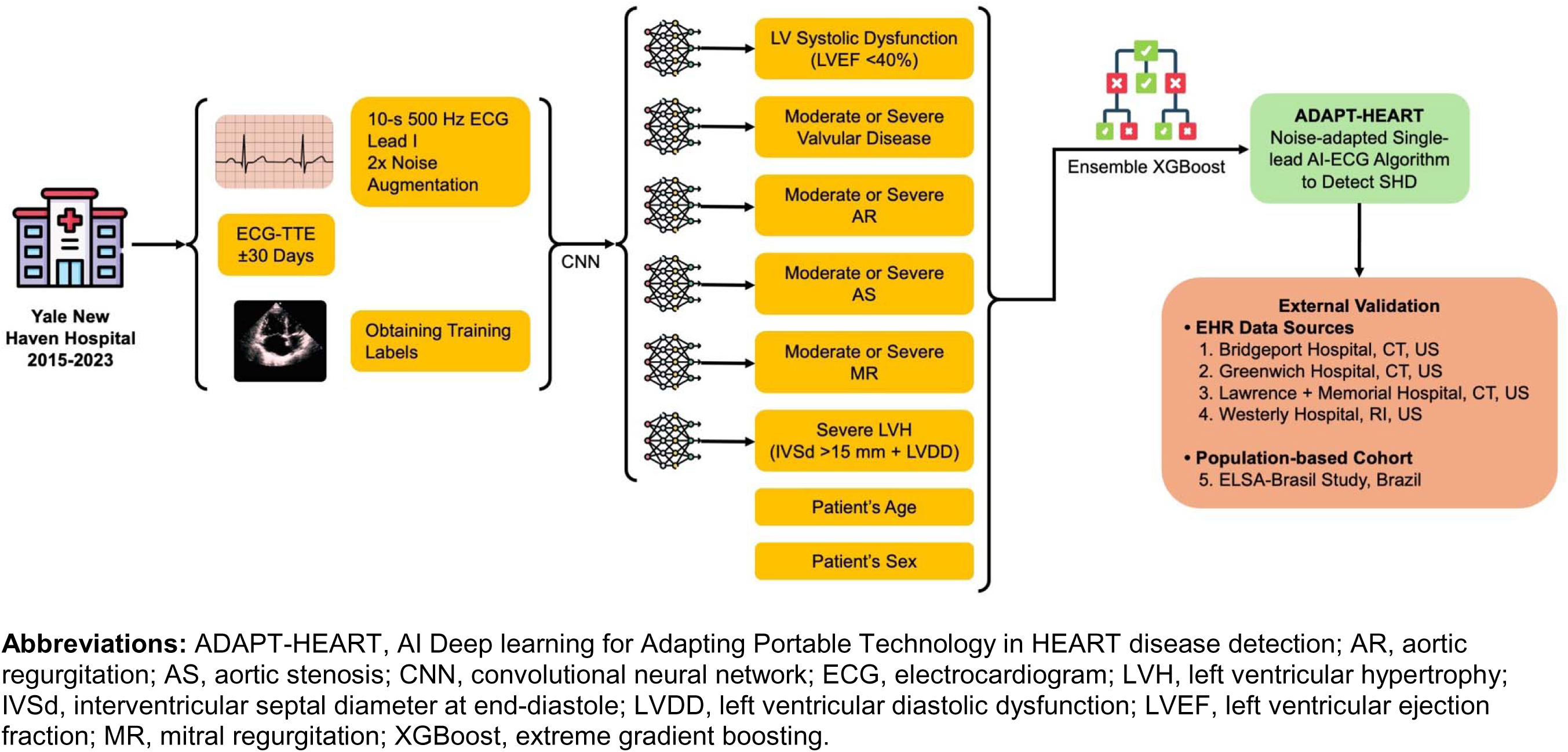
Model Development for Detecting Multiple Structural Heart Diseases.

### Study Outcome

The primary study outcome was the model’s discrimination for detecting the presence of composite SHD, measured by the area under the receiver operating characteristics curve (AUROC). The exploratory outcome was the model’s performance for predicting new-onset SHD in individuals without baseline SHD, measured by the hazard ratio (HR) of high- vs. low-risk groups. We defined the risk groups by the model output probability using the threshold for optimizing sensitivity at 90%. For the predictive assessment, incident SHD was characterized as TTE-defined SHD, heart failure hospitalization based on diagnosis codes, or left-sided valve replacement or repair based on procedure codes in YNHH and hospital-based external validation sites. In the UKB, due to the absence of serial imaging, incident SHD was characterized by hospitalization for heart failure, or the first recorded diagnosis code of left-sided valvular disease or procedure codes for left-sided valve replacement or repair (**Table S1**).

### Statistical Analysis

Continuous and categorical variables were reported as medians and interquartile ranges (IQRs) and numbers and percentages, respectively. The model’s performance was presented using AUROC, along with sensitivity, specificity, positive predictive value (PPV), negative predictive value (NPV), and F1 score of the model across the thresholds and specifically for the threshold corresponding to a sensitivity of 90% using the internal validation set. The 95% confidence intervals (CI) for AUROC were calculated using bootstrapping with 1000 iterations. We computed 95% CI for sensitivity, specificity, PPV, and NPV using the standard error formula for proportion. To quantify the model calibration, we calculated the Brier score, which is the mean squared difference between the predicted probabilities and the actual outcomes, ranging from 0 to 1, with values closer to 0 representing good calibration.^22^

For the predictive assessment, we fit age- and sex-adjusted Cox proportional hazard models with incident SHD as the dependent variable and the risk groups defined by the model output probability using the threshold for optimizing sensitivity at 90% as the key independent variable. These risk groups comprised those with a high probability of SHD but without SHD, false positives, compared with those with a low probability of SHD and without SHD, true negatives. We further adjusted the Cox models for baseline hypertension and diabetes mellitus. Additionally, we accounted for the competing risk of death using the Fine-Gray subdistribution hazard model.^23^

All statistical analyses were executed using Python 3.11.2, and R version 4.2.0. Statistical tests were two-sided with the significance level set at 0.05. The Yale Institutional Review Board approved the study protocol and waived the need for informed consent as the study involves analyzing pre-existing data. Patients who opted out of research studies were not included in the study. Participants from ELSA-Brasil and the UKB provided informed consent, and their de-identified data were analyzed in this study.^19,20^ We used the UK Biobank Resource under Application Number 71033.

## RESULTS

### Study Population

The model was developed in 266,740 ECGs with paired TTE data from 99,205 unique patients with a median age of 66 [54-77] years. This included 49,947 (50.3%) women, 13,503 (14.0%) non-Hispanic Black, and 7,832 (8.1%) Hispanic individuals (**Table 1**). In the development set, 60,096 (22.5%) ECGs were linked to a TTE with SHD, including 25,552 (9.6%) with LVSD, 42,989 (16.1%) with moderate or severe left-sided valvular disease, and 1,004 (0.4%) with sLVH (**Table 1**). The training set included 261,228 ECGs from 93,693 unique patients, while the validation and held-out test sets included 5,512 and 11,023 ECGs, respectively, with one ECG drawn per person (**Figure S1**, **Table S2**). For external validation, we included 65,988 patients from four community hospitals and 3,014 participants from the ELSA-Brasil with diverse demographic backgrounds (**Table 1**, **Figure S1**). The prevalence of SHD varied from 20.2%-27.1% in hospital-based external validation sites compared with 2.9% in ELSA-Brasil (**Table 1**).

**Table 1.**
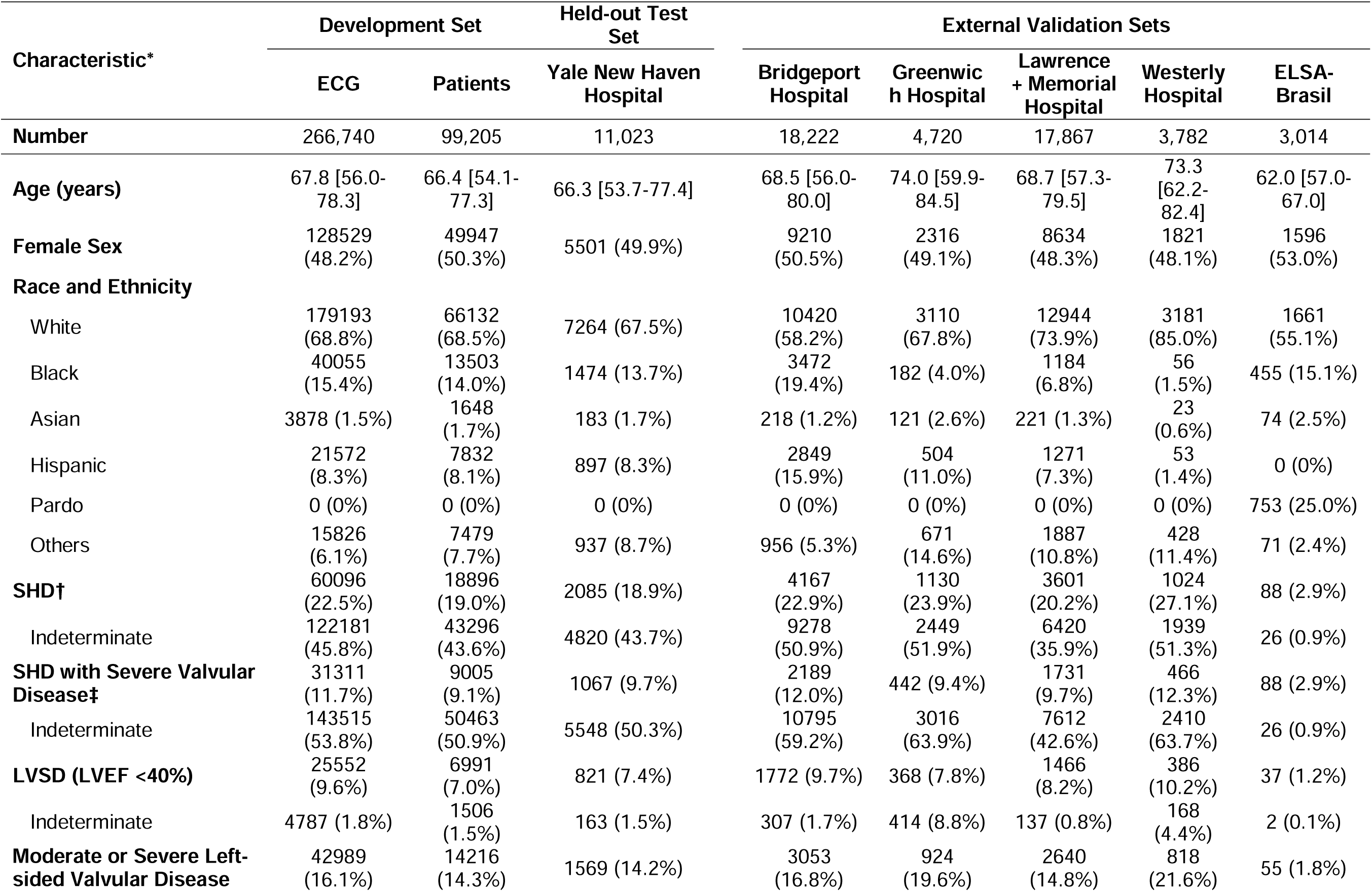
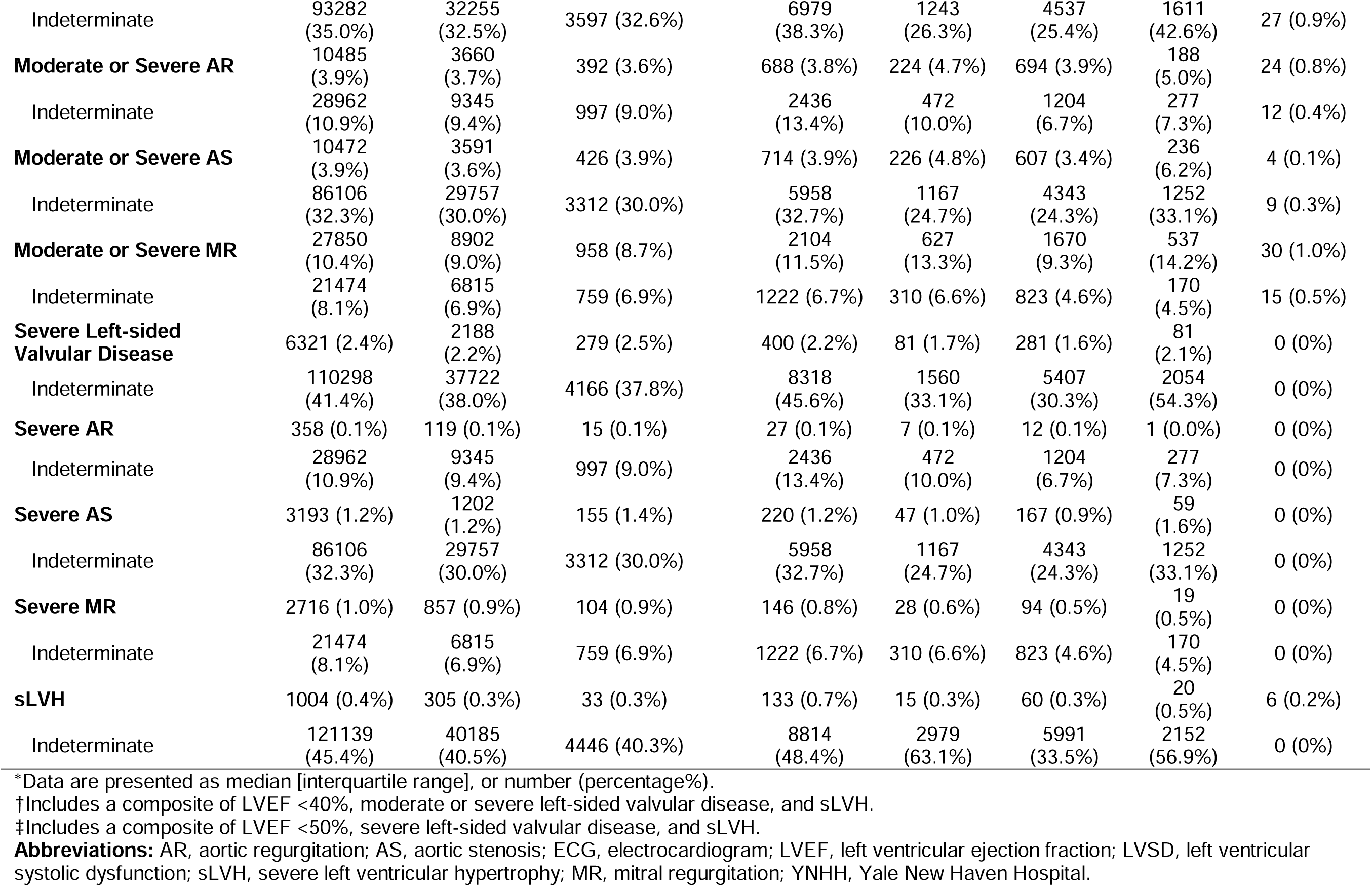
Demographics and Prevalence of Structural Heart Disease in the Development and External Validation Set at Patient- and ECG-level.

| Characteristic* | Development Set |  | Held-out Test Set | External Validation Sets |  |  |  |  |
| --- | --- | --- | --- | --- | --- | --- | --- | --- |
|  | ECG | Patients | Yale New Haven Hospital | Bridgeport Hospital | Greenwich Hospital | Lawrence + Memorial Hospital | Westerly Hospital | ELSA-Brasil |
| **Number** | 266,740 | 99,205 | 11,023 | 18,222 | 4,720 | 17,867 | 3,782 | 3,014 |
| **Age (years)** | 67.8 [56.0-78.3] | 66.4 [54.1-77.3] | 66.3 [53.7-77.4] | 68.5 [56.0-80.0] | 74.0 [59.9-84.5] | 68.7 [57.3-79.5] | 73.3 [62.2-82.4] | 62.0 [57.0-67.0] |
| **Female Sex** | 128529 (48.2%) | 49947 (50.3%) | 5501 (49.9%) | 9210 (50.5%) | 2316 (49.1%) | 8634 (48.3%) | 1821 (48.1%) | 1596 (53.0%) |
| **Race and Ethnicity** |  |  |  |  |  |  |  |  |
| White | 179193 (68.8%) | 66132 (68.5%) | 7264 (67.5%) | 10420 (58.2%) | 3110 (67.8%) | 12944 (73.9%) | 3181 (85.0%) | 1661 (55.1%) |
| Black | 40055 (15.4%) | 13503 (14.0%) | 1474 (13.7%) | 3472 (19.4%) | 182 (4.0%) | 1184 (6.8%) | 56 (1.5%) | 455 (15.1%) |
| Asian | 3878 (1.5%) | 1648 (1.7%) | 183 (1.7%) | 218 (1.2%) | 121 (2.6%) | 221 (1.3%) | 23 (0.6%) | 74 (2.5%) |
| Hispanic | 21572 (8.3%) | 7832 (8.1%) | 897 (8.3%) | 2849 (15.9%) | 504 (11.0%) | 1271 (7.3%) | 53 (1.4%) | 0 (0%) |
| Pardo | 0 (0%) | 0 (0%) | 0 (0%) | 0 (0%) | 0 (0%) | 0 (0%) | 0 (0%) | 753 (25.0%) |
| Others | 15826 (6.1%) | 7479 (7.7%) | 937 (8.7%) | 956 (5.3%) | 671 (14.6%) | 1887 (10.8%) | 428 (11.4%) | 71 (2.4%) |
| **SHD†** | 60096 (22.5%) | 18896 (19.0%) | 2085 (18.9%) | 4167 (22.9%) | 1130 (23.9%) | 3601 (20.2%) | 1024 (27.1%) | 88 (2.9%) |
| Indeterminate | 122181 (45.8%) | 43296 (43.6%) | 4820 (43.7%) | 9278 (50.9%) | 2449 (51.9%) | 6420 (35.9%) | 1939 (51.3%) | 26 (0.9%) |
| **SHD with Severe Valvular Disease‡** | 31311 (11.7%) | 9005 (9.1%) | 1067 (9.7%) | 2189 (12.0%) | 442 (9.4%) | 1731 (9.7%) | 466 (12.3%) | 88 (2.9%) |
| Indeterminate | 143515 (53.8%) | 50463 (50.9%) | 5548 (50.3%) | 10795 (59.2%) | 3016 (63.9%) | 7612 (42.6%) | 2410 (63.7%) | 26 (0.9%) |
| **LVSD (LVEF <40%)** | 25552 (9.6%) | 6991 (7.0%) | 821 (7.4%) | 1772 (9.7%) | 368 (7.8%) | 1466 (8.2%) | 386 (10.2%) | 37 (1.2%) |
| Indeterminate | 4787 (1.8%) | 1506 (1.5%) | 163 (1.5%) | 307 (1.7%) | 414 (8.8%) | 137 (0.8%) | 168 (4.4%) | 2 (0.1%) |
| **Moderate or Severe Left-sided Valvular Disease** | 42989 (16.1%) | 14216 (14.3%) | 1569 (14.2%) | 3053 (16.8%) | 924 (19.6%) | 2640 (14.8%) | 818 (21.6%) | 55 (1.8%) |
| Indeterminate | 93282<br>(35.0%) | 32255<br>(32.5%) | 3597 (32.6%) | 6979<br>(38.3%) | 1243<br>(26.3%) | 4537<br>(25.4%) | 1611<br>(42.6%) | 27 (0.9%) |
| **Moderate or Severe AR** | 10485<br>(3.9%) | 3660<br>(3.7%) | 392 (3.6%) | 688 (3.8%) | 224 (4.7%) | 694 (3.9%) | 188<br>(5.0%) | 24 (0.8%) |
| Indeterminate | 28962<br>(10.9%) | 9345<br>(9.4%) | 997 (9.0%) | 2436<br>(13.4%) | 472<br>(10.0%) | 1204<br>(6.7%) | 277<br>(7.3%) | 12 (0.4%) |
| **Moderate or Severe AS** | 10472<br>(3.9%) | 3591<br>(3.6%) | 426 (3.9%) | 714 (3.9%) | 226 (4.8%) | 607 (3.4%) | 236<br>(6.2%) | 4 (0.1%) |
| Indeterminate | 86106<br>(32.3%) | 29757<br>(30.0%) | 3312 (30.0%) | 5958<br>(32.7%) | 1167<br>(24.7%) | 4343<br>(24.3%) | 1252<br>(33.1%) | 9 (0.3%) |
| **Moderate or Severe MR** | 27850<br>(10.4%) | 8902<br>(9.0%) | 958 (8.7%) | 2104<br>(11.5%) | 627<br>(13.3%) | 1670<br>(9.3%) | 537<br>(14.2%) | 30 (1.0%) |
| Indeterminate | 21474<br>(8.1%) | 6815<br>(6.9%) | 759 (6.9%) | 1222 (6.7%) | 310 (6.6%) | 823 (4.6%) | 170<br>(4.5%) | 15 (0.5%) |
| **Severe Left-sided**<br>**Valvular Disease** | 6321 (2.4%) | 2188<br>(2.2%) | 279 (2.5%) | 400 (2.2%) | 81 (1.7%) | 281 (1.6%) | 81<br>(2.1%) | 0 (0%) |
| Indeterminate | 110298<br>(41.4%) | 37722<br>(38.0%) | 4166 (37.8%) | 8318<br>(45.6%) | 1560<br>(33.1%) | 5407<br>(30.3%) | 2054<br>(54.3%) | 0 (0%) |
| **Severe AR** | 358 (0.1%) | 119 (0.1%) | 15 (0.1%) | 27 (0.1%) | 7 (0.1%) | 12 (0.1%) | 1 (0.0%) | 0 (0%) |
| Indeterminate | 28962<br>(10.9%) | 9345<br>(9.4%) | 997 (9.0%) | 2436<br>(13.4%) | 472<br>(10.0%) | 1204<br>(6.7%) | 277<br>(7.3%) | 0 (0%) |
| **Severe AS** | 3193 (1.2%) | 1202<br>(1.2%) | 155 (1.4%) | 220 (1.2%) | 47 (1.0%) | 167 (0.9%) | 59<br>(1.6%) | 0 (0%) |
| Indeterminate | 86106<br>(32.3%) | 29757<br>(30.0%) | 3312 (30.0%) | 5958<br>(32.7%) | 1167<br>(24.7%) | 4343<br>(24.3%) | 1252<br>(33.1%) | 0 (0%) |
| **Severe MR** | 2716 (1.0%) | 857 (0.9%) | 104 (0.9%) | 146 (0.8%) | 28 (0.6%) | 94 (0.5%) | 19<br>(0.5%) | 0 (0%) |
| Indeterminate | 21474<br>(8.1%) | 6815<br>(6.9%) | 759 (6.9%) | 1222 (6.7%) | 310 (6.6%) | 823 (4.6%) | 170<br>(4.5%) | 0 (0%) |
| **sLVH** | 1004 (0.4%) | 305 (0.3%) | 33 (0.3%) | 133 (0.7%) | 15 (0.3%) | 60 (0.3%) | 20<br>(0.5%) | 6 (0.2%) |
| Indeterminate | 121139<br>(45.4%) | 40185<br>(40.5%) | 4446 (40.3%) | 8814<br>(48.4%) | 2979<br>(63.1%) | 5991<br>(33.5%) | 2152<br>(56.9%) | 0 (0%) |

\*Data are presented as median [interquartile range], or number (percentage%).
†Includes a composite of LVEF <40%, moderate or severe left-sided valvular disease, and sLVH.
‡Includes a composite of LVEF <50%, severe left-sided valvular disease, and sLVH.
**Abbreviations:** AR, aortic regurgitation; AS, aortic stenosis; ECG, electrocardiogram; LVEF, left ventricular ejection fraction; LVSD, left ventricular systolic dysfunction; sLVH, severe left ventricular hypertrophy; MR, mitral regurgitation; YNHH, Yale New Haven Hospital.

### Detection of Structural Heart Disease

ADAPT-HEART demonstrated an AUROC of 0.879 (95% CI, 0.870-0.888) for detecting SHD on a single-lead ECG in the YNHH held-out test set (**Figure 2A**, **Table S3**). The model was well-calibrated for detecting the primary study outcome with a Brier score of 0.083. ADAPT-HEART had higher discrimination (AUROC, 0.910 [95% CI, 0.900-0.920]) when the composite SHD label included severe, instead of moderate or severe, left-sided valvular disease (**Figure 2B**). The model’s probability threshold corresponding to a sensitivity of 90% for predicting SHD in the internal validation set was 0.190. At this probability threshold, in the held-out test set, ADAPT-HEART had a sensitivity of 90.9% (95% CI, 90.2-91.6), specificity of 61.8% (95% CI, 60.6-63.0), PPV of 54.6% (95% CI, 53.4-55.9), and NPV of 93.1% (95% CI, 92.4-93.7) for detecting SHD. A higher probability threshold, aimed at optimizing the F1 score, had higher specificity and PPV (**Figure 3**, **Table S4**).

**Figure 2.**
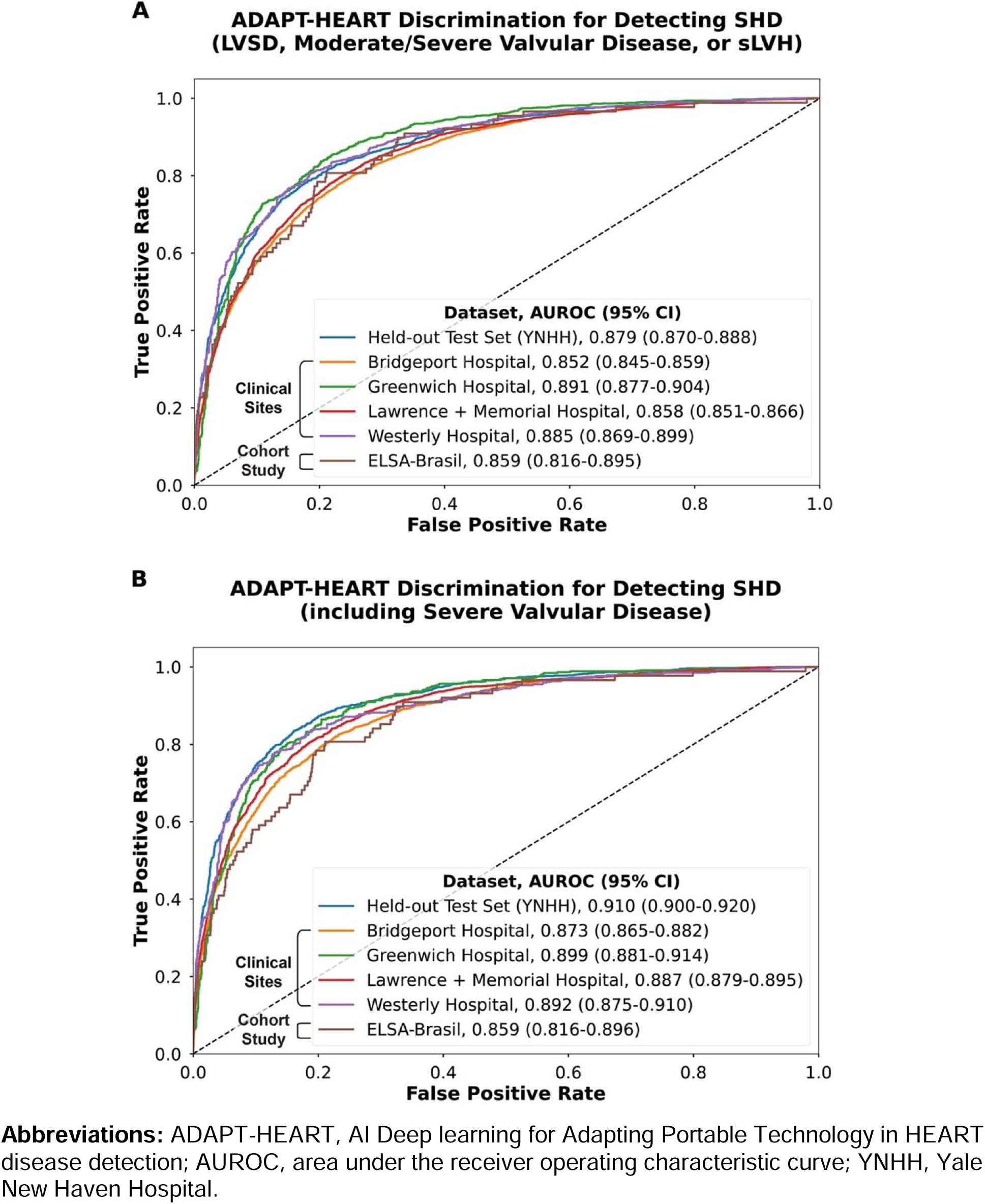
Receiver Operating Characteristic Curves of the ADAPT-HEART for Detecting (A) the Primary and (B) the Secondary Outcome Structural Heart Disease in the Held-out Test Set and External Validation Cohorts.

**Figure 3.**
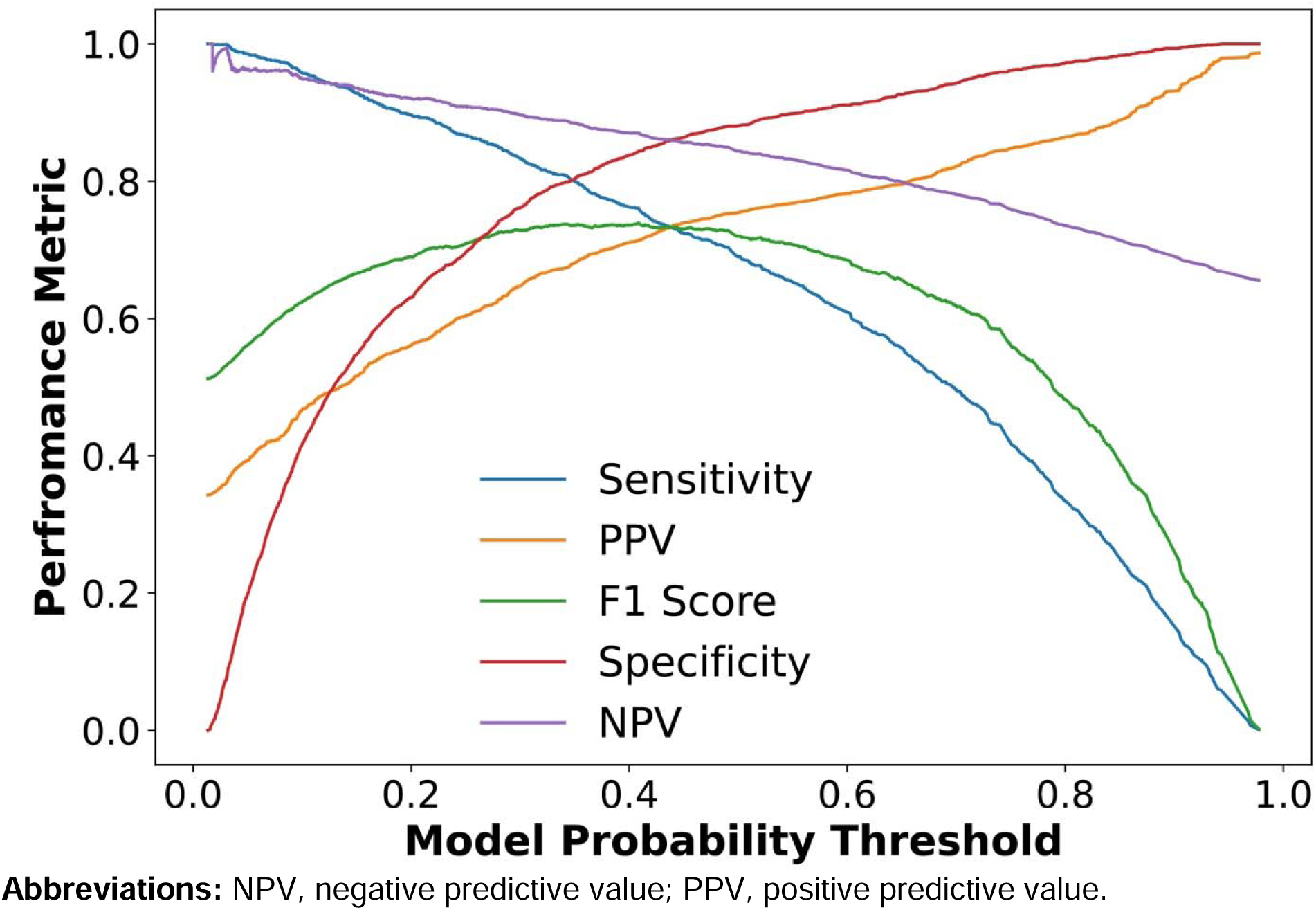
Model’s Performance Measures Across Thresholds in the Held-out Test Set.

ADAPT-HEART performed consistently across external validation cohorts, with the AUROC varying from 0.852 (95% CI, 0.845-0.859) at Bridgeport Hospital to 0.891 (95% CI, 0.877-0.904) with good calibration across clinical sites (**Figure 2**, **Table S3**). In the prospective ELSA-Brasil that had protocolized paired ECGs and TTEs, ADAPT-HEART retained high discrimination and good calibration with an AUROC of 0.859 (95% CI, 0.816-0.895) (**Figure 2**). The model’s performance was comparable across key demographic subgroups of age, sex, and race and ethnicity in the held-out test set and external validation sites (**Figure 4**, **Tables S5-10**). Individual label-specific models demonstrated consistent performance in the YNHH held-out test set and external validation sites (**Tables S11-16**, **Figure S2**)

**Figure 4.**
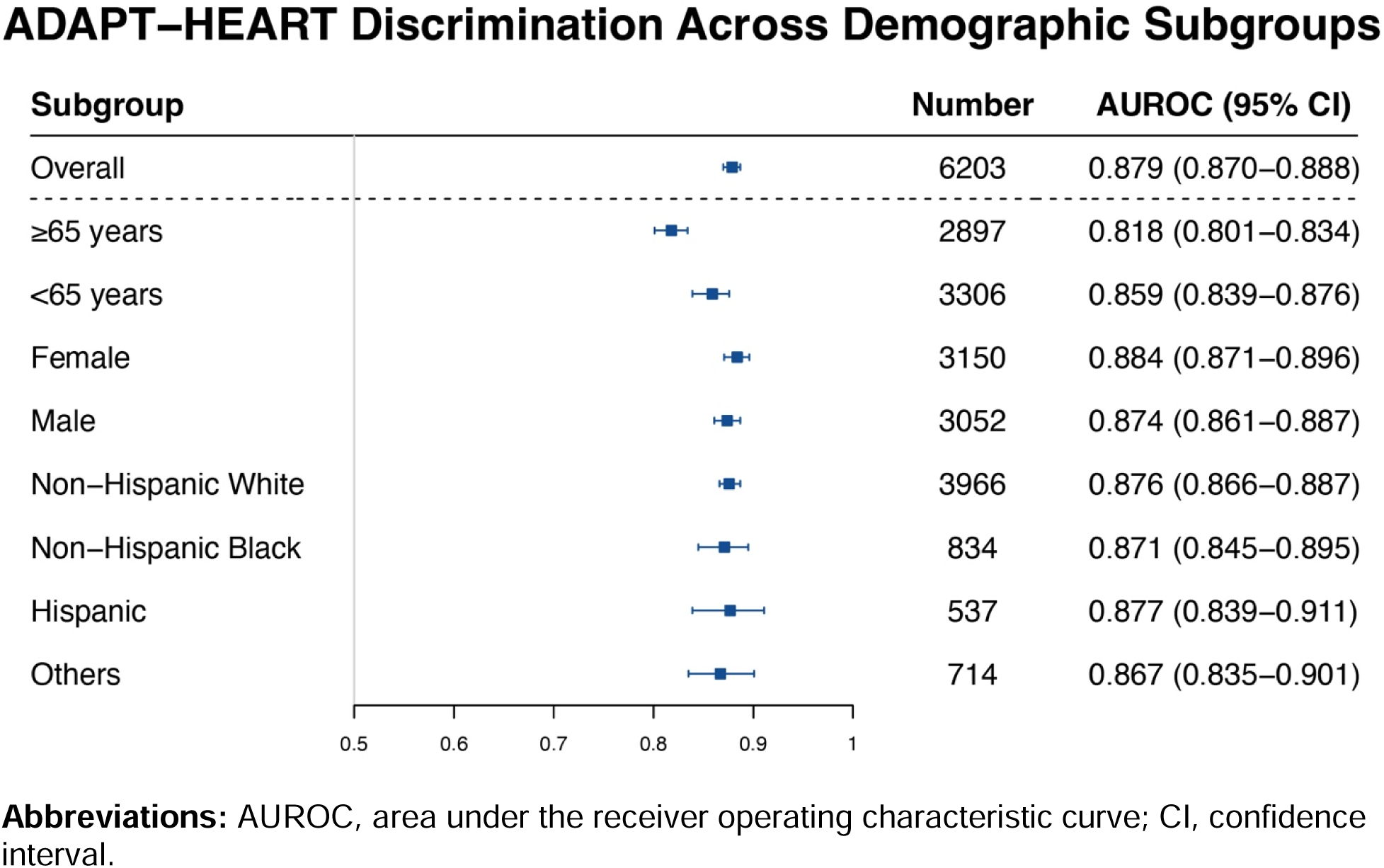
Model’s Performance for Detecting Structural Heart Disease Across Key Demographic Subgroups in the Held-out Test Set.

### Predictive Performance of ADAPT-HEART

For the predictive assessment, we identified 127,547 individuals from YNHH, 105,992 from hospital-based external validation sites, and 41,800 from the UKB (**Table S17**). Over a median follow-up of 4.0 [1.7-6.4] years, 5,353 (4.2%) patients developed new-onset SHD in YNHH compared with 5.6% to 8.1% across hospital-based external validation sites with the median follow-up ranging from 2.4 to 4.7 years (**Table S17**). In the UKB, 413 (1.0%) participants had new-onset SHD over a median follow-up of 3.0 [2.1-4.5] years. A screen-positive single-lead ECG defined by ADAPT-HEART was associated with a 4-fold higher hazard of incident SHD in YNHH (adjusted HR, 4.03; 95% CI, 3.71-4.37), with a consistently elevated risk across hospital-based cohorts (**Table 2**). In the UKB, a positive AI-ECG portended a nearly 3-fold hazard (HR: 2.82; 95% CI, 2.13-3.74) for incident SHD, with a modified definition that did not include serial imaging. The risk of SHD was consistently observed with a positive AI-ECG screen, independent of additional adjustment for cardiovascular risk factors of hypertension and diabetes mellitus, and also the competing risk of death (**Table 2**).

**Table 2.**
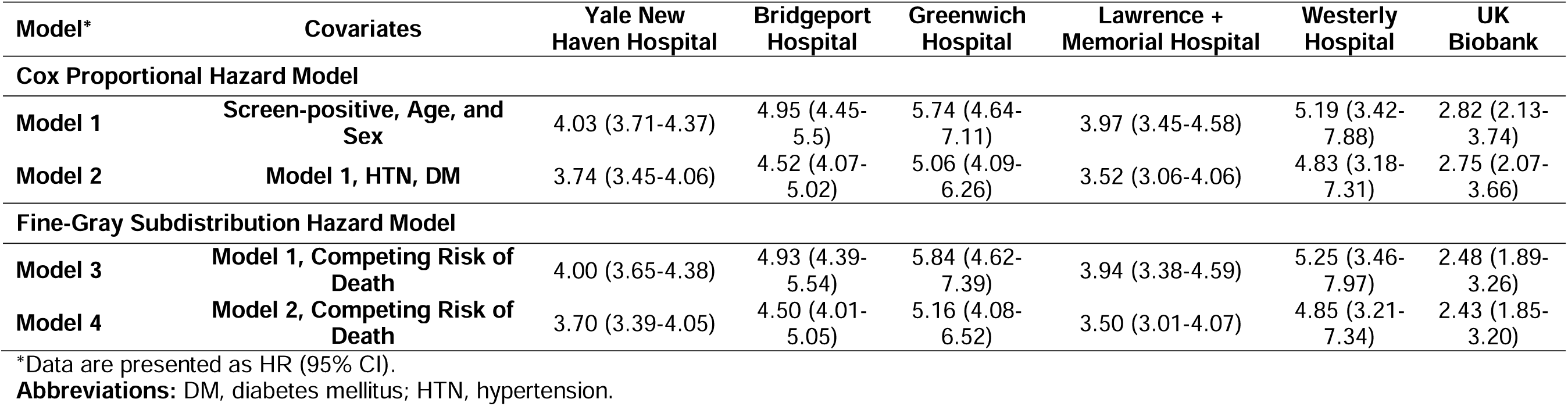
Hazard Ratio of Screen-positive ECG Based on ADAPT-HEART for Predicting Incident Structural Heart Disease in Yale New Haven Hospital, Hospital-based External Validation Sites, and the UK Biobank.

| Model* | Covariates | Yale New Haven Hospital | Bridgeport Hospital | Greenwich Hospital | Lawrence + Memorial Hospital | Westerly Hospital | UK Biobank |
| --- | --- | --- | --- | --- | --- | --- | --- |
| **Cox Proportional Hazard Model** |  |  |  |  |  |  |  |
| **Model 1** | **Screen-positive, Age, and Sex** | 4.03 (3.71-4.37) | 4.95 (4.45-5.5) | 5.74 (4.64-7.11) | 3.97 (3.45-4.58) | 5.19 (3.42-7.88) | 2.82 (2.13-3.74) |
| **Model 2** | **Model 1, HTN, DM** | 3.74 (3.45-4.06) | 4.52 (4.07-5.02) | 5.06 (4.09-6.26) | 3.52 (3.06-4.06) | 4.83 (3.18-7.31) | 2.75 (2.07-3.66) |
| **Fine-Gray Subdistribution Hazard Model** |  |  |  |  |  |  |  |
| **Model 3** | **Model 1, Competing Risk of Death** | 4.00 (3.65-4.38) | 4.93 (4.39-5.54) | 5.84 (4.62-7.39) | 3.94 (3.38-4.59) | 5.25 (3.46-7.97) | 2.48 (1.89-3.26) |
| **Model 4** | **Model 2, Competing Risk of Death** | 3.70 (3.39-4.05) | 4.50 (4.01-5.05) | 5.16 (4.08-6.52) | 3.50 (3.01-4.07) | 4.85 (3.21-7.34) | 2.43 (1.85-3.20) |

\*Data are presented as HR (95% CI).
**Abbreviations:** DM, diabetes mellitus; HTN, hypertension.

## DISCUSSION

We developed and externally validated ADAPT-HEART, a noise-adapted deep learning algorithm that detects and predicts SHD from single-lead ECGs obtainable on portable and wearable devices, using a large and diverse population. Our novel noise-adapted strategy for single-lead ECGs demonstrates robust discrimination and calibration for detecting a composite of multiple clinically significant SHDs, including LVSD, left-sided valvular disease, and sLVH, using age, sex, and a single-lead ECG recording as the only inputs. ADAPT-HEART performed comparably across external validation cohorts and key demographic subgroups. Depending on the prevalence of SHD in the target screening population, the screening strategy informed by ADAPT-HEART requires 2-12 individuals to undergo TTE to find one case with any SHD. Additionally, individuals without baseline SHD but with high ADAPT-HEART probability were three to six times more likely to develop SHD.

ADAPT-HEART builds upon the current literature by providing a scalable strategy for SHD screening in the community using single-lead ECGs acquired on portable and wearable devices.^16,24–26^ The advantages of using single-lead ECGs compared with 12-lead ECGs for SHD screening are two-fold; first, the feasibility of obtaining single-lead ECGs on portable devices expands the scale of screening from healthcare settings to broader communities, and second, it allows targeting individuals regardless of their access to healthcare. Additionally, ADAPT-HEART leverages our previous work to retain robust performance despite the noisy acquisition of real-world portable ECGs.^14,15,17^ We also employed an ensemble learning strategy that enables the learning of granular ECG signatures for individual SHDs, resulting in superior performance compared with a single multi-label CNN model for detecting the composite SHD label.^16^ Thus, the rapidly increasing number of portable and wearable device users, combined with the ADAPT-HEART development, significantly enhances SHD screening strategies in the communities.^12,13,27^

In addition to the diagnostic value of ADAPT-HEART, we demonstrated its role as a biomarker for predicting incident SHD using single-lead ECGs. Thus, ADAPT-HEART not only enables the identification of individuals with asymptomatic SHD but also enables risk stratification for those without SHD. This approach can be particularly valuable to risk stratify asymptomatic individuals at risk for SHD, such as those with hypertension and ischemic heart disease, and older adults.^28^ The lack of an evidence-based protocolized follow-up for such individuals underscores the need for precision tools such as ADAPT-HEART to provide personalized care to at-risk people. Additionally, the predictive significance of ADAPT-HEART for new-onset SHD may serve as its clinical explainability, where a high model probability in those without baseline SHD can indicate electrical signatures of SHD in the subclinical stages.

Leveraging ADAPT-HEART and single-lead ECGs for SHD screening in the community can transform cardiovascular care by enabling early detection and timely treatment. Beyond the use case of ADAPT-HEART for SHD screening among the growing number of portable and wearable device users, it can also enhance screening for individuals without such devices or those less likely to access healthcare settings.^29^ Public health promotion programs, such as hypertension screening in barbershops and pharmacies or cancer screening in churches, can be adapted to establish SHD screening programs using portable ECGs.^30,31^ These community outreach efforts would allow the screening of a large number of individuals for SHD using a limited number of portable devices, offering an efficient strategy to identify individuals who may benefit from advanced cardiac imaging.^32,33^ Furthermore, the proposed screening strategy can help prioritize echocardiographic studies in underserved areas, including low- and middle-income countries.^34,35^ Therefore, our study establishes new frameworks to improve SHD diagnosis at scale and serves as a tool that has the potential to reduce disparities in SHD care.

Our findings should be interpreted in the light of the following limitations. First, ADAPT-HEART was developed using pairs of ECG-TTE among individuals who underwent these tests within 30 days of each other, representing a selective population. However, the model performed consistently across four hospitals and a population-based cohort study, indicating generalizability to distinct settings. Second, hospitals in the same health system as the development set were part of model validation sets. Nonetheless, we excluded patients from the external validation sites whose data were used for model development to ensure independence of development and external validation populations. Furthermore, the model generalized well to the population-based cohort of ELSA-Brasil, suggesting its robustness and external validity outside the Yale New Haven Health System. Third, we developed the model using lead I extracted from 12-lead ECGs rather than portable-acquired ECGs. However, the lack of a large and rigorous data source where portable ECGs are paired with advanced cardiac imaging data precludes the development of such a model using portable ECGs. Additionally, single-lead AI-ECG algorithms that were developed using clinical 12-lead ECGs have been shown to retain performance while deployed to real-world smartwatch ECGs.^36^ We also adopted a noising strategy for model development to enhance the model’s resilience to noise in real-world settings.^17^

## CONCLUSION

We propose a novel model that detects and predicts a range of SHDs from noisy single-lead ECGs obtainable on portable/wearable devices, providing a scalable strategy for community-based screening and risk stratification for SHD.

## Supporting information

Supplemental Content

## Acknowledgments

None.

## Data Availability Statement

The data from the Yale New Haven Health System represent protected health information. To protect patient privacy, the Yale Institutional Review Board does not allow sharing of these data. Data from the Brazilian Longitudinal Study of Adult Health and the UK Biobank are available for research to licensed users.

