## Supplemental Content for "Development and Multinational Validation of an Ensemble Deep Learning Algorithm for Detecting and Predicting Structural Heart Disease Using Noisy Single-lead Electrocardiograms"

### **SUPPLEMENTAL METHODS**

**Signal Preprocessing**

We isolated raw voltage data for lead I⎯representing the standard lead captured by portable devices⎯of 12-lead electrocardiogram (ECG) obtained as standard 10-second 12-lead ECGs at a sampling frequency of 500 Hz or 250 Hz. We then employed a standard signal preprocessing strategy to extract the signal data, which involved median pass filtering and scaling for each lead to harmonize the included ECGs. Median filtering was conducted by subtracting a one-second median filter from the acquired signals to eliminate baseline drift. ECG signals were also divided by 1000 to scale them to millivolts. ECGs acquired at 250 Hz were up-sampled to 500 Hz to align with the predominant frequency across data sources.

**Noising Strategy**

To construct algorithms resilient to the noise introduced during the acquisition of ECGs obtained from portable devices, we artificially incorporated noises into the model development process as described previously.^1^ We augmented ECGs in the training set using random Gaussian noises, while we tested the model on clean ECGs that were not noised. We isolated four distinct noises from a 5-minute random Gaussian noise within four frequency ranges of 3-12 Hz, 12-50 Hz, 50-100 Hz, and 100-150 Hz, with each of which modeling a specific type of real-world noise. The noise with a frequency range of 3-12 Hz reflects the motion artifact noises attributable to tremors, 50-100 Hz accounts for the electrode contact noise, and 12-50 Hz and 100-150 Hz reflect the lower and higher-frequency muscle noises, respectively.^2,3^ Each ECG in the training set was included two times with different random noises for model training. This augmentation involved a random type of noise and a random signal-to-noise ratio (SNR). For this purpose, we first randomly selected one of the four abovementioned distinct random Gaussian noises. Finally, the selected noise was introduced to the ECG waveform with a random SNR ranging from 0.5 to 1.25, representing a heavy and a light burden of noise in ECGs, respectively.

**Convolutional Neural Network Training**

The employed convolutional neural network (CNN) architecture comprised an input layer with dimensions of (5000, 1, 1), representing a 10-second, 500 Hz, lead I ECG.^1^ The input layer was followed by seven 2-dimensional convolutional layers, progressively increasing the number of filters from 16 to 64 while incorporating varying kernel sizes (7x1, 5x1, and 3x1) to capture different levels of feature abstraction. A batch normalization layer, a rectified linear unit (ReLU) activation layer, and a 2-dimensional max-pooling layer with different pool sizes (2x1 and 4x1) followed each convolutional layer. Next, the output of the 7^th^ convolutional layer was used as the input for a fully connected network that included two dense layers. Each dense layer was followed by a batch normalization layer, a ReLU activation layer, and a dropout layer with a rate of 0.5. Finally, the model output was a dense layer with a single class and a sigmoid activation to generate the output probability of the label. The loss function was adjusted by calculating model weights using a class re-weighting approach to ensure that the learning is not impacted by the differential prevalence of positive and negative labels.^4^

The left ventricular systolic dysfunction (LVSD) model was trained first and the weights from the optimal epoch were transferred to initialize the training for the models for valvular disease and severe left ventricular hypertrophy (sLVH) labels. Each model training was stopped early if the validation loss did not improve for five consecutive epochs. After the completion of training for each model, the optimal epoch was selected based on low loss and high area under the receiver operating characteristic curve (AUROC) in the internal validation set. All models were trained using the Adam optimizer on the Keras framework in TensorFlow 2.9.1 and Python 3.9.0.

### **Table S1. Diagnosis and Procedure Codes for Defining Structural Heart Diseases and Study Covariates in the Predictive Assessment Across Hospital-based Sites and the UK Biobank**

| **Condition** | **Coding System** | **Codes** |
| --- | --- | --- |
| **Hospital-based Sites** | | |
| Heart Failure | ICD-10-CM | I110, I130, I132, I50, I500, I501, I509, Z9581, I0981 |
| Aortic Valve Replacement or Repair | CPT4 | 33361, 33362, 33363, 33364, 33365, 33366, 33367, 33368, 33369, 33370, 33390, 33391, 33404, 33405, 33406, 33410, 33440, 33411, 33412, 33413, 33414, 33415, 33416, 33417 |
|  | ICD-10-PCS | Contains 'aortic valve', 'opn aortic valvuloplasty' in description |
| Mitral Valve Replacement or Repair | CPT4 | 33418, 33419, 33420, 33422, 33425, 33426, 33427, 33430, 33440 |
|  | ICD-10-PCS | Contains ‘mitral’ in description |
| Type 2 Diabetes Mellitus | ICD-10-CM | E11, E110, E111, E112, E113, E114, E115, E116, E117, E118, E119, O241 |
| Hypertension | ICD-10-CM | I10, I11, I110, I119, I12, I120, I129, I13, I130, I131, I132, I139, I674, O10, O100, O101, O102, O103, O109, O11 |
| **UK Biobank** | | |
| Heart Failure | ICD-10 | I110, I130, I132, I50, I500, I501, I509 |
|  | ICD-9 | 428, 4280, 4281, 4289 |
| Aortic Valve Disease | ICD-10 | I06, I060, I061, I062, I068, I069, I08, I080, I082, I083, I088, I089, I35, I350, I351, I352, I358, I359, Z952, Z953, Z954 |
|  | ICD-9 | 395, 3950, 3951, 3952, 3959, 396, 3969, 4241, V422, V433 |
| Aortic Valve Replacement or Repair | OPCS-4 | K26, K261, K262, K263, K264, K265, K268, K269, K29, K291, K292, K293, K294, K295, K298, K299, K30, K302, K308, K309, K31, K312, K318, K319, K32, K322, K328, K329, K35, K352, K355, K358, K359 |
|  | OPCS-3 | 311, 3112, 3115, 312, 3122, 3125, 313, 3132, 3135, 314, 3142, 3145 |
| Mitral Valve Disease | ICD-10 | I05, I050, I051, I052, I058, I059, I08, I080, I081, I083, I088, I089, I34, I340, I341, I342, I348, I349, Z952, Z953, Z954 |
|  | ICD-9 | 394, 3940, 3941, 3942, 3949, 396, 3969, 4240, V422, V433 |
| Mitral Valve Replacement or Repair | OPCS-4 | K25, K251 K252, K253, K254, K255, K258, K259, K29, K291, K292, K293, K294, K295, K298, K299, K30, K301, K308, K309, K31, K311, K318, K319, K32, K321, K328, K329, K34, K341, K343, K348, K349, K35, K351, K355, K358, K359 |
|  | OPCS-3 | 311, 3111, 3115, 312, 3121, 3125, 313, 3131, 3135, 314, 3141, 3145 |
| Type 2 Diabetes Mellitus | ICD-10 | E11, E110, E111, E112, E113, E114, E115, E116, E117, E118, E119, O241, O242, O243, O249 |
|  | ICD-9 | 250, 2500, 25000, 25009, 2501, 25010, 25019, 2502, 25020, 25029, 2503, 2504, 2505, 2506, 2507, 2509, 25090, 25099, 6480 |
| Hypertension | ICD-10 | I10, I11, I110, I119, I12, I120, I129, I13, I130, I131, I132, I139, I674, O10, O100, O101, O102, O103, O109, O11 |
|  | ICD-9 | 401, 4010, 4011, 4019, 402, 4020, 4021, 4029, 403, 4030, 4031, 4039, 404, 4040, 4041, 4049, 6420, 6422, 6427, 6429 |

**Abbreviations:** CPT4, Current Procedural Terminology 4; ICD-9 and ICD-10, International Classification of Diseases, Ninth and Tenth Revisions; ICD-10-CM, International Classification of Diseases, Tenth Revision, Clinical Modification; ICD-10-PCS, and procedure codes were recorded as the International Classification of Diseases, Tenth Revision, Procedure Coding System; OPCS-3 and OPCS-4, Office of Population Censuses and Surveys Classification of Interventions and Procedures, versions 3 and 4.

### **Table S2. Demographics and Prevalence of Structural Heart Disease Across Train, Internal Validation, and Held-out Test Sets**

| **Characteristic*** | **Train Set (ECG Level)** | **Train Set (Patient Level)** | **Internal Validation Set** | **Held-out Test Set** |
| --- | --- | --- | --- | --- |
| **Number** | 261,228 | 93,693 | 5,512 | 11,023 |
| **Age (years)** | 67.8 [56.1-78.3] | 66.4 [54.1-77.3] | 66.5 [54.1-77.4] | 66.3 [53.7-77.4] |
| **Female Sex** | 125735 (48.1%) | 47153 (50.3%) | 2794 (50.7%) | 5501 (49.9%) |
| **Race and Ethnicity** |  |  |  |  |
| White | 175531 (68.8%) | 62470 (68.5%) | 3662 (68.5%) | 7264 (67.5%) |
| Black | 39298 (15.4%) | 12746 (14.0%) | 757 (14.2%) | 1474 (13.7%) |
| Asian | 3788 (1.5%) | 1558 (1.7%) | 90 (1.7%) | 183 (1.7%) |
| Hispanic | 21159 (8.3%) | 7419 (8.1%) | 413 (7.7%) | 897 (8.3%) |
| Others | 15399 (6.0%) | 7052 (7.7%) | 427 (8.0%) | 937 (8.7%) |
| **SHD†** | 59005 (22.6%) | 17805 (19.0%) | 1091 (19.8%) | 2085 (18.9%) |
| Indeterminate | 119837 (45.9%) | 40952 (43.7%) | 2344 (42.5%) | 4820 (43.7%) |
| **SHD with Severe Valvular Disease‡** | 30786 (11.8%) | 8480 (9.1%) | 525 (9.5%) | 1067 (9.7%) |
| Indeterminate | 140753 (53.9%) | 47701 (50.9%) | 2762 (50.1%) | 5548 (50.3%) |
| **LVSD (LVEF <40%)** | 25162 (9.6%) | 6601 (7.0%) | 390 (7.1%) | 821 (7.4%) |
| Indeterminate | 4705 (1.8%) | 1424 (1.5%) | 82 (1.5%) | 163 (1.5%) |
| **Moderate or Severe Left-sided Valvular Disease** | 42170 (16.1%) | 13397 (14.3%) | 819 (14.9%) | 1569 (14.2%) |
| Indeterminate | 91537 (35.0%) | 30510 (32.6%) | 1745 (31.7%) | 3597 (32.6%) |
| **Moderate or Severe AR** | 10271 (3.9%) | 3446 (3.7%) | 214 (3.9%) | 392 (3.6%) |
| Indeterminate | 28423 (10.9%) | 8806 (9.4%) | 539 (9.8%) | 997 (9.0%) |
| **Moderate or Severe AS** | 10270 (3.9%) | 3389 (3.6%) | 202 (3.7%) | 426 (3.9%) |
| Indeterminate | 84492 (32.3%) | 28143 (30.0%) | 1614 (29.3%) | 3312 (30.0%) |
| **Moderate or Severe MR** | 27347 (10.5%) | 8399 (9.0%) | 503 (9.1%) | 958 (8.7%) |
| Indeterminate | 21087 (8.1%) | 6428 (6.9%) | 387 (7.0%) | 759 (6.9%) |
| **Severe Left-sided Valvular Disease** | 6193 (2.4%) | 2060 (2.2%) | 128 (2.3%) | 279 (2.5%) |
| Indeterminate | 108234 (41.4%) | 35658 (38.1%) | 2064 (37.4%) | 4166 (37.8%) |
| **Severe AR** | 348 (0.1%) | 109 (0.1%) | 10 (0.2%) | 15 (0.1%) |
| Indeterminate | 28423 (10.9%) | 8806 (9.4%) | 539 (9.8%) | 997 (9.0%) |
| **Severe AS** | 3123 (1.2%) | 1132 (1.2%) | 70 (1.3%) | 155 (1.4%) |
| Indeterminate | 84492 (32.3%) | 28143 (30.0%) | 1614 (29.3%) | 3312 (30.0%) |
| **Severe MR** | 2672 (1.0%) | 813 (0.9%) | 44 (0.8%) | 104 (0.9%) |
| Indeterminate | 21087 (8.1%) | 6428 (6.9%) | 387 (7.0%) | 759 (6.9%) |
| **sLVH** | 975 (0.4%) | 276 (0.3%) | 29 (0.5%) | 33 (0.3%) |
| Indeterminate | 118906 (45.5%) | 37952 (40.5%) | 2233 (40.5%) | 4446 (40.3%) |

*Data are presented as median [interquartile range], or number (percentage%).

†Includes a composite of LVEF <40%, moderate or severe left-sided valvular disease, and sLVH.

‡Includes a composite of LVEF <50%, severe left-sided valvular disease, and sLVH.

**Abbreviations:** AR, aortic regurgitation; AS, aortic stenosis; ECG, electrocardiogram; LVEF, left ventricular ejection fraction; LVSD, left ventricular systolic dysfunction; sLVH, severe left ventricular hypertrophy; MR, mitral regurgitation.

### **Table S3. Model’s Performance Measures for Detecting Structural Heart Disease in the Held-out Test Set and Across External Validation Sites Using the Threshold for Optimizing Sensitivity at 90%**

| **Subgroup** | **Total Number** | **Diagnostic OR** | **AUROC** | **Brier Score** | **F1 Score** | **Prevalence** | **Sensitivity** | **Specificity** | **PPV** | **NPV** |
| --- | --- | --- | --- | --- | --- | --- | --- | --- | --- | --- |
| **Held-out Test Set** | |  |  |  |  |  |  |  |  |  |
| Yale New Haven Hospital | 6203 | 16.1 (13.7-18.9) | 0.879 (0.870-0.888) | 0.083 | 0.682 | 33.6% | 90.9% (90.2-91.6) | 61.8% (60.6-63.0) | 54.6% (53.4-55.9) | 93.1% (92.4-93.7) |
| **External Validation Sites** | |  |  |  |  |  |  |  |  |  |
| Bridgeport Hospital | 8944 | 14.0 (12.2-15.9) | 0.852 (0.845-0.859) | 0.115 | 0.745 | 46.6% | 93.4% (92.9-93.9) | 49.9% (48.9-50.9) | 61.9% (60.9-62.9) | 89.7% (89.0-90.3) |
| Greenwich Hospital | 2271 | 26.4 (19.4-36.0) | 0.891 (0.877-0.904) | 0.096 | 0.791 | 49.8% | 95.6% (94.7-96.4) | 54.2% (52.1-56.2) | 67.4% (65.4-69.3) | 92.5% (91.4-93.6) |
| Lawrence + Memorial Hospital | 11447 | 14.7 (12.8-16.9) | 0.858 (0.851-0.866) | 0.087 | 0.624 | 31.5% | 93.4% (93.0-93.9) | 51.3% (50.4-52.2) | 46.8% (45.9-47.8) | 94.5% (94.0-94.9) |
| Westerly Hospital | 1843 | 19.6 (14.0-27.3) | 0.885 (0.869-0.899) | 0.112 | 0.803 | 55.6% | 95.9% (95.0-96.8) | 46.2% (43.9-48.4) | 69.0% (66.9-71.1) | 90.0% (88.6-91.4) |
| ELSA-Brasil | 2988 | 11.8 (6.8-20.4) | 0.859 (0.816-0.895) | 0.054 | 0.149 | 2.9% | 81.8% (80.4-83.2) | 72.4% (70.8-74.0) | 8.2% (7.3-9.2) | 99.2% (98.9-99.6) |

**Abbreviations:** AUROC, area under the receiver operating characteristic curve; NPV, negative predictive value; OR, odds ratio; PPV, positive predictive value.

### **Table S4. Performance Measures of ADAPT-HEART for Detecting Structural Heart Disease Across Different Thresholds in the Held-out Test Set at Yale New Haven Hospital**

| **Performance Metric** | **YNHH Held-out Test Set** | **Greenwich Hospital** | **Lawrence + Memorial Hospital** | **Bridgeport Hospital** | **Westerly Hospital** | **ELSA-Brasil** |
| --- | --- | --- | --- | --- | --- | --- |
| **Optimal Sensitivity** | |  |  |  |  |  |
| Sensitivity | 90.9% (90.2-91.6) | 95.6% (94.7-96.4) | 93.4% (93.0-93.9) | 93.4% (92.9-93.9) | 95.9% (95.0-96.8) | 81.8% (80.4-83.2) |
| Specificity | 61.8% (60.6-63.0) | 54.2% (52.1-56.2) | 51.3% (50.4-52.2) | 49.9% (48.9-50.9) | 46.2% (43.9-48.4) | 72.4% (70.8-74.0) |
| PPV | 54.6% (53.4-55.9) | 67.4% (65.4-69.3) | 46.8% (45.9-47.8) | 61.9% (60.9-62.9) | 69.0% (66.9-71.1) | 8.2% (7.3-9.2) |
| NPV | 93.1% (92.4-93.7) | 92.5% (91.4-93.6) | 94.5% (94.0-94.9) | 89.7% (89.0-90.3) | 90.0% (88.6-91.4) | 99.2% (98.9-99.6) |
| F1 Score | 0.682 | 0.791 | 0.624 | 0.745 | 0.803 | 0.149 |
| **Optimal Youden's Index** | |  |  |  |  |  |
| Sensitivity | 80.7% (79.7-81.7) | 90.0% (88.8-91.2) | 85.5% (84.8-86.1) | 85.3% (84.5-86.0) | 89.4% (87.9-90.8) | 53.4% (51.6-55.2) |
| Specificity | 79.6% (78.6-80.6) | 70.5% (68.6-72.3) | 69.5% (68.6-70.3) | 67.4% (66.4-68.4) | 67.3% (65.1-69.4) | 92.0% (91.0-92.9) |
| PPV | 66.7% (65.5-67.9) | 75.1% (73.3-76.9) | 56.2% (55.3-57.1) | 69.5% (68.6-70.5) | 77.3% (75.4-79.3) | 16.8% (15.4-18.1) |
| NPV | 89.1% (88.3-89.8) | 87.7% (86.3-89.0) | 91.2% (90.7-91.8) | 84.0% (83.2-84.7) | 83.5% (81.8-85.2) | 98.5% (98.0-98.9) |
| F1 Score | 0.73 | 0.819 | 0.678 | 0.766 | 0.829 | 0.256 |
| **Optimal F1 Score** | |  |  |  |  |  |
| Sensitivity | 76.0% (74.9-77.0) | 87.2% (85.8-88.5) | 81.0% (80.3-81.7) | 81.0% (80.2-81.8) | 84.9% (83.2-86.5) | 42.0% (40.3-43.8) |
| Specificity | 84.0% (83.1-85.0) | 75.4% (73.6-77.1) | 75.1% (74.3-75.9) | 73.2% (72.3-74.1) | 74.4% (72.4-76.4) | 95.0% (94.2-95.8) |
| PPV | 70.7% (69.5-71.8) | 77.8% (76.1-79.5) | 59.9% (59.0-60.8) | 72.5% (71.6-73.4) | 80.5% (78.7-82.3) | 20.3% (18.9-21.8) |
| NPV | 87.4% (86.5-88.2) | 85.6% (84.1-87.0) | 89.6% (89.0-90.2) | 81.5% (80.7-82.3) | 79.7% (77.9-81.5) | 98.2% (97.7-98.7) |
| F1 Score | 0.733 | 0.822 | 0.689 | 0.765 | 0.826 | 0.274 |
| **Optimal Specificity** |  |  |  |  |  |  |
| Sensitivity | 65.7% (64.5-66.9) | 76.8% (75.1-78.5) | 70.5% (69.6-71.3) | 70.9% (69.9-71.8) | 77.1% (75.1-79.0) | 29.5% (27.9-31.2) |
| Specificity | 90.0% (89.3-90.8) | 84.4% (82.9-85.9) | 84.0% (83.4-84.7) | 82.7% (81.9-83.5) | 84.4% (82.7-86.0) | 98.0% (97.5-98.5) |
| PPV | 77.0% (75.9-78.0) | 83.0% (81.4-84.5) | 66.9% (66.1-67.8) | 78.1% (77.2-79.0) | 86.0% (84.5-87.6) | 31.0% (29.3-32.6) |
| NPV | 83.8% (82.9-84.8) | 78.6% (76.9-80.3) | 86.1% (85.5-86.7) | 76.5% (75.6-77.4) | 74.6% (72.6-76.6) | 97.9% (97.3-98.4) |
| F1 Score | 0.709 | 0.798 | 0.687 | 0.743 | 0.813 | 0.302 |

**Abbreviations:** NPV, negative predictive value; PPV, positive predictive value; YNHH, Yale New Haven Hospital.

### **Table S5. Model’s Performance Measures for Detecting Structural Heart Disease Across Key Demographic Subgroups in the Internal Held-out Test Set at Yale New Haven Hospital**

| **Subgroup** | **Total Number** | **Diagnostic OR** | **AUROC** | **F1 Score** | **Prevalence** | **Sensitivity** | **Specificity** | **PPV** | **NPV** |
| --- | --- | --- | --- | --- | --- | --- | --- | --- | --- |
| **Overall** | 6203 | 16.1 (13.7-18.9) | 0.879 (0.870-0.888) | 0.686 | 33.6% | 90.5% (89.8-91.2) | 62.8% (61.6-64.0) | 55.2% (54.0-56.5) | 92.9% (92.3-93.5) |
| **≥65 years** | 2897 | 9.4 (6.8-13.1) | 0.818 (0.801-0.834) | 0.73 | 53.2% | 97.2% (96.6-97.8) | 21.2% (19.7-22.7) | 58.4% (56.6-60.2) | 87.0% (85.8-88.2) |
| **<65 years** | 3306 | 12.5 (10.1-15.4) | 0.859 (0.839-0.876) | 0.558 | 16.5% | 71.5% (70.0-73.0) | 83.3% (82.0-84.5) | 45.7% (44.0-47.4) | 93.7% (92.9-94.5) |
| **Female** | 3150 | 18.5 (14.7-23.4) | 0.884 (0.871-0.896) | 0.677 | 30.8% | 90.6% (89.6-91.6) | 65.7% (64.1-67.4) | 54.1% (52.3-55.8) | 94.0% (93.2-94.9) |
| **Male** | 3052 | 13.9 (11.2-17.3) | 0.874 (0.861-0.887) | 0.694 | 36.5% | 90.4% (89.3-91.4) | 59.6% (57.9-61.3) | 56.3% (54.5-58.0) | 91.5% (90.5-92.5) |
| **Non-Hispanic White** | 3966 | 16.9 (13.7-20.9) | 0.876 (0.866-0.887) | 0.701 | 37.4% | 92.7% (91.9-93.5) | 57.0% (55.5-58.6) | 56.4% (54.8-57.9) | 92.9% (92.1-93.7) |
| **Non-Hispanic Black** | 834 | 14.2 (9.5-21.4) | 0.871 (0.845-0.895) | 0.673 | 31.5% | 87.8% (85.6-90.1) | 66.4% (63.2-69.6) | 54.6% (51.2-58.0) | 92.2% (90.4-94.0) |
| **Hispanic** | 537 | 15.5 (9.3-25.7) | 0.877 (0.839-0.911) | 0.651 | 25.5% | 83.9% (80.8-87.0) | 74.8% (71.1-78.4) | 53.2% (49.0-57.5) | 93.1% (91.0-95.3) |
| **Others** | 714 | 11.7 (7.5-18.3) | 0.867 (0.835-0.901) | 0.578 | 21.0% | 80.7% (77.8-83.6) | 73.8% (70.5-77.0) | 45.0% (41.3-48.6) | 93.5% (91.7-95.3) |

**Abbreviations:** AUROC, area under the receiver operating characteristic curve; NPV, negative predictive value; OR, odds ratio; PPV, positive predictive value.

### **Table S6. Model’s Performance Measures for Detecting Structural Heart Disease Across Key Demographic Subgroups in Bridgeport Hospital**

| **Subgroup** | **Total Number** | **Diagnostic OR** | **AUROC** | **F1 Score** | **Prevalence** | **Sensitivity** | **Specificity** | **PPV** | **NPV** |
| --- | --- | --- | --- | --- | --- | --- | --- | --- | --- |
| **Overall** | 8944 | 14.0 (12.2-15.9) | 0.852 (0.845-0.859) | 0.745 | 46.6% | 93.2% (92.7-93.7) | 50.4% (49.4-51.4) | 62.1% (61.1-63.1) | 89.5% (88.8-90.1) |
| **≥65 years** | 5139 | 9.2 (6.9-12.3) | 0.797 (0.785-0.809) | 0.772 | 60.3% | 98.1% (97.7-98.5) | 15.2% (14.2-16.2) | 63.7% (62.4-65.0) | 84.0% (83.0-85.0) |
| **<65 years** | 3805 | 12.4 (10.4-14.7) | 0.854 (0.840-0.868) | 0.662 | 28.1% | 79.0% (77.7-80.3) | 76.7% (75.3-78.0) | 56.9% (55.3-58.5) | 90.4% (89.4-91.3) |
| **Female** | 4528 | 12.0 (10.0-14.4) | 0.836 (0.824-0.848) | 0.727 | 44.7% | 92.3% (91.5-93.1) | 50.1% (48.6-51.5) | 59.9% (58.5-61.3) | 88.9% (88.0-89.8) |
| **Male** | 4416 | 16.4 (13.4-19.9) | 0.867 (0.857-0.878) | 0.764 | 48.5% | 94.1% (93.4-94.8) | 50.8% (49.3-52.2) | 64.3% (62.9-65.7) | 90.1% (89.2-91.0) |
| **Non-Hispanic White** | 5010 | 13.2 (10.9-16.0) | 0.841 (0.830-0.852) | 0.77 | 53.3% | 95.0% (94.4-95.6) | 41.1% (39.7-42.5) | 64.8% (63.5-66.1) | 87.8% (86.9-88.7) |
| **Non-Hispanic Black** | 1754 | 11.7 (8.9-15.4) | 0.844 (0.824-0.863) | 0.714 | 41.5% | 90.4% (89.0-91.8) | 55.5% (53.1-57.8) | 59.0% (56.7-61.3) | 89.0% (87.6-90.5) |
| **Hispanic** | 1355 | 15.1 (11.0-20.8) | 0.866 (0.847-0.885) | 0.72 | 38.0% | 89.7% (88.1-91.3) | 63.5% (60.9-66.0) | 60.1% (57.5-62.7) | 91.0% (89.4-92.5) |
| **Others** | 700 | 13.9 (8.5-22.6) | 0.853 (0.821-0.883) | 0.632 | 29.0% | 89.7% (87.4-91.9) | 61.6% (58.0-65.2) | 48.8% (45.1-52.5) | 93.6% (91.8-95.4) |

**Abbreviations:** AUROC, area under the receiver operating characteristic curve; NPV, negative predictive value; OR, odds ratio; PPV, positive predictive value.

### **Table S7. Model’s Performance Measures for Detecting Structural Heart Disease Across Key Demographic Subgroups in Greenwich Hospital**

| **Subgroup** | **Total Number** | **Diagnostic OR** | **AUROC** | **F1 Score** | **Prevalence** | **Sensitivity** | **Specificity** | **PPV** | **NPV** |
| --- | --- | --- | --- | --- | --- | --- | --- | --- | --- |
| **Overall** | 2271 | 26.4 (19.4-36.0) | 0.891 (0.877-0.904) | 0.792 | 49.8% | 95.7% (94.8-96.5) | 54.5% (52.5-56.6) | 67.6% (65.6-69.5) | 92.7% (91.6-93.8) |
| **≥65 years** | 1440 | 8.2 (4.6-14.4) | 0.821 (0.795-0.843) | 0.815 | 67.2% | 98.3% (97.7-99.0) | 12.1% (10.4-13.8) | 69.6% (67.3-72.0) | 78.1% (75.9-80.2) |
| **<65 years** | 831 | 21.2 (13.7-32.8) | 0.886 (0.852-0.915) | 0.653 | 19.5% | 79.6% (76.9-82.4) | 84.5% (82.0-86.9) | 55.4% (52.0-58.7) | 94.5% (92.9-96.0) |
| **Female** | 1139 | 26.1 (16.6-41.2) | 0.892 (0.873-0.910) | 0.772 | 47.4% | 95.9% (94.8-97.1) | 52.6% (49.7-55.5) | 64.6% (61.8-67.4) | 93.5% (92.0-94.9) |
| **Male** | 1132 | 27.2 (17.9-41.5) | 0.892 (0.872-0.910) | 0.811 | 52.1% | 95.4% (94.2-96.6) | 56.6% (53.8-59.5) | 70.6% (67.9-73.2) | 91.9% (90.3-93.5) |
| **Non-Hispanic White** | 1440 | 28.5 (18.4-44.2) | 0.875 (0.857-0.894) | 0.825 | 58.8% | 97.2% (96.3-98.0) | 45.5% (42.9-48.0) | 71.7% (69.4-74.1) | 91.8% (90.4-93.3) |
| **Non-Hispanic Black** | 77 | 7.1 (2.5-20.1) | 0.853 (0.758-0.928) | 0.738 | 49.4% | 81.6% (72.9-90.2) | 61.5% (50.7-72.4) | 67.4% (56.9-77.9) | 77.4% (68.1-86.8) |
| **Hispanic** | 235 | 26.3 (11.7-59.2) | 0.886 (0.836-0.926) | 0.785 | 41.3% | 91.8% (88.2-95.3) | 70.3% (64.4-76.1) | 68.5% (62.5-74.4) | 92.4% (89.0-95.8) |
| **Others** | 465 | 22.6 (11.1-46.2) | 0.901 (0.864-0.931) | 0.635 | 26.7% | 92.7% (90.4-95.1) | 63.9% (59.6-68.3) | 48.3% (43.8-52.9) | 96.0% (94.3-97.8) |

**Abbreviations:** AUROC, area under the receiver operating characteristic curve; NPV, negative predictive value; OR, odds ratio; PPV, positive predictive value.

### **Table S8. Model’s Performance Measures for Detecting Structural Heart Disease Across Key Demographic Subgroups in Lawrence + Memorial Hospital**

| **Subgroup** | **Total Number** | **Diagnostic OR** | **AUROC** | **F1 Score** | **Prevalence** | **Sensitivity** | **Specificity** | **PPV** | **NPV** |
| --- | --- | --- | --- | --- | --- | --- | --- | --- | --- |
| **Overall** | 11447 | 14.7 (12.8-16.9) | 0.858 (0.851-0.866) | 0.626 | 31.5% | 93.2% (92.7-93.6) | 51.9% (51.0-52.8) | 47.1% (46.2-48.0) | 94.3% (93.9-94.7) |
| **≥65 years** | 6165 | 10.5 (7.9-14.1) | 0.805 (0.794-0.816) | 0.658 | 45.5% | 98.2% (97.9-98.5) | 16.3% (15.4-17.2) | 49.5% (48.3-50.8) | 91.5% (90.8-92.2) |
| **<65 years** | 5282 | 11.2 (9.4-13.4) | 0.843 (0.826-0.858) | 0.508 | 15.0% | 75.4% (74.2-76.6) | 78.5% (77.4-79.7) | 38.3% (37.0-39.6) | 94.8% (94.2-95.4) |
| **Female** | 5634 | 13.8 (11.4-16.7) | 0.848 (0.837-0.858) | 0.616 | 30.5% | 92.5% (91.8-93.2) | 52.9% (51.6-54.2) | 46.2% (44.9-47.5) | 94.1% (93.5-94.7) |
| **Male** | 5813 | 15.7 (12.9-19.1) | 0.868 (0.858-0.878) | 0.634 | 32.4% | 93.8% (93.2-94.4) | 51.0% (49.7-52.3) | 47.9% (46.6-49.1) | 94.5% (93.9-95.1) |
| **Non-Hispanic White** | 8085 | 14.8 (12.5-17.6) | 0.854 (0.845-0.862) | 0.643 | 34.8% | 94.4% (93.9-94.9) | 46.9% (45.9-48.0) | 48.7% (47.6-49.7) | 94.0% (93.5-94.5) |
| **Non-Hispanic Black** | 776 | 14.0 (8.6-22.9) | 0.857 (0.823-0.885) | 0.606 | 27.2% | 90.5% (88.5-92.6) | 59.5% (56.0-62.9) | 45.5% (42.0-49.0) | 94.4% (92.8-96.0) |
| **Hispanic** | 882 | 16.3 (9.9-26.9) | 0.876 (0.845-0.905) | 0.546 | 19.5% | 89.0% (86.9-91.0) | 66.9% (63.8-70.0) | 39.4% (36.2-42.7) | 96.2% (94.9-97.4) |
| **Others** | 1488 | 10.3 (7.4-14.4) | 0.833 (0.807-0.858) | 0.552 | 23.3% | 86.7% (85.0-88.5) | 61.2% (58.7-63.7) | 40.5% (38.0-43.0) | 93.8% (92.6-95.0) |

**Abbreviations:** AUROC, area under the receiver operating characteristic curve; NPV, negative predictive value; OR, odds ratio; PPV, positive predictive value.

### **Table S9. Model’s Performance Measures for Detecting Structural Heart Disease Across Key Demographic Subgroups in Westerly Hospital**

| **Subgroup** | **Total Number** | **Diagnostic OR** | **AUROC** | **F1 Score** | **Prevalence** | **Sensitivity** | **Specificity** | **PPV** | **NPV** |
| --- | --- | --- | --- | --- | --- | --- | --- | --- | --- |
| **Overall** | 1843 | 19.6 (14.0-27.3) | 0.885 (0.869-0.899) | 0.803 | 55.6% | 95.7% (94.8-96.6) | 46.8% (44.5-49.0) | 69.2% (67.1-71.3) | 89.7% (88.3-91.1) |
| **≥65 years** | 1282 | 27.9 (12.7-61.0) | 0.851 (0.828-0.872) | 0.832 | 67.6% | 99.2% (98.7-99.7) | 18.5% (16.4-20.6) | 71.7% (69.2-74.2) | 91.7% (90.2-93.2) |
| **<65 years** | 561 | 10.3 (6.7-15.9) | 0.857 (0.821-0.891) | 0.644 | 28.2% | 76.6% (73.1-80.1) | 75.9% (72.4-79.5) | 55.5% (51.4-59.6) | 89.2% (86.6-91.8) |
| **Female** | 895 | 17.6 (11.0-28.1) | 0.878 (0.854-0.900) | 0.788 | 53.6% | 95.4% (94.0-96.8) | 45.8% (42.5-49.0) | 67.1% (64.0-70.1) | 89.6% (87.6-91.6) |
| **Male** | 948 | 21.7 (13.6-34.7) | 0.891 (0.869-0.911) | 0.818 | 57.4% | 96.0% (94.7-97.2) | 47.8% (44.6-51.0) | 71.2% (68.3-74.1) | 89.8% (87.8-91.7) |
| **Non-Hispanic White** | 1527 | 19.2 (13.4-27.7) | 0.880 (0.862-0.898) | 0.825 | 60.2% | 96.0% (95.0-97.0) | 44.6% (42.2-47.1) | 72.4% (70.2-74.7) | 88.0% (86.4-89.6) |
| **Non-Hispanic Black** | 23 | 10.5 (1.0-108.6) | 0.825 (0.607-0.973) | 0.666 | 34.8% | 87.5% (74.0-101.0) | 60.0% (40.0-80.0) | 53.8% (33.5-74.2) | 90.0% (77.7-102.3) |
| **Hispanic** | 28 | 7.2 (0.7-72.7) | 0.894 (0.719-1.000) | 0.5 | 21.4% | 83.3% (69.5-97.1) | 59.1% (40.9-77.3) | 35.7% (18.0-53.5) | 92.9% (83.3-102.4) |
| **Others** | 242 | 16.2 (6.2-42.1) | 0.887 (0.838-0.931) | 0.653 | 34.3% | 94.0% (91.0-97.0) | 50.9% (44.6-57.2) | 50.0% (43.7-56.3) | 94.2% (91.2-97.1) |

**Abbreviations:** AUROC, area under the receiver operating characteristic curve; NPV, negative predictive value; OR, odds ratio; PPV, positive predictive value.

### **Table S10. Model’s Performance Measures for Detecting Structural Heart Disease Across Key Demographic Subgroups in the ELSA-Brasil Cohort**

| **Subgroup** | **Total Number** | **Diagnostic OR** | **AUROC** | **F1 Score** | **Prevalence** | **Sensitivity** | **Specificity** | **PPV** | **NPV** |
| --- | --- | --- | --- | --- | --- | --- | --- | --- | --- |
| **Overall** | 2988 | 11.8 (6.8-20.4) | 0.859 (0.816-0.895) | 0.149 | 2.9% | 81.8% (80.4-83.2) | 72.4% (70.8-74.0) | 8.2% (7.3-9.2) | 99.2% (98.9-99.6) |
| **≥65 years** | 1087 | 5.3 (2.4-11.9) | 0.801 (0.733-0.865) | 0.131 | 4.5% | 85.7% (83.6-87.8) | 46.9% (44.0-49.9) | 7.1% (5.6-8.6) | 98.6% (97.9-99.3) |
| **<65 years** | 1901 | 21.5 (10.1-45.8) | 0.879 (0.807-0.938) | 0.188 | 2.1% | 76.9% (75.0-78.8) | 86.6% (85.0-88.1) | 10.7% (9.3-12.1) | 99.4% (99.1-99.8) |
| **Female** | 1584 | 7.8 (3.6-16.8) | 0.853 (0.804-0.898) | 0.106 | 2.1% | 72.7% (70.5-74.9) | 74.4% (72.3-76.6) | 5.7% (4.6-6.8) | 99.2% (98.8-99.7) |
| **Male** | 1404 | 16.0 (7.2-35.8) | 0.857 (0.796-0.913) | 0.189 | 3.9% | 87.3% (85.5-89.0) | 70.1% (67.7-72.4) | 10.6% (9.0-12.2) | 99.3% (98.8-99.7) |
| **White** | 1644 | 13.8 (5.7-33.1) | 0.872 (0.823-0.914) | 0.124 | 2.4% | 84.6% (82.9-86.4) | 71.5% (69.3-73.6) | 6.7% (5.5-7.9) | 99.5% (99.1-99.8) |
| **Black** | 451 | 12.1 (4.1-35.9) | 0.887 (0.825-0.941) | 0.24 | 5.5% | 84.0% (80.6-87.4) | 69.7% (65.5-74.0) | 14.0% (10.8-17.2) | 98.7% (97.6-99.7) |
| **Pardo** | 748 | 9.5 (3.7-24.4) | 0.802 (0.687-0.904) | 0.164 | 3.1% | 73.9% (70.8-77.1) | 77.0% (73.9-80.0) | 9.2% (7.2-11.3) | 98.9% (98.2-99.7) |
| **Others** | 145 | - | 0.924 | 0.041 | 0.7% | 100.0% | 67.4% | 2.1% | 100.0% |

**Abbreviations:** AUROC, area under the receiver operating characteristic curve; NPV, negative predictive value; OR, odds ratio; PPV, positive predictive value.

### **Table S11. Performance Measures of the Label-specific Convolutional Neural Network Model for Detecting Left Ventricular Systolic Dysfunction in the Held-out Test Set and Across External Validation Sites**

| **Subgroup** | **Total Number** | **Diagnostic OR** | **AUROC** | **F1 Score** | **Prevalence** | **Sensitivity** | **Specificity** | **PPV** | **NPV** |
| --- | --- | --- | --- | --- | --- | --- | --- | --- | --- |
| **Held-out Test Set** |  |  |  |  |  |  |  |  |  |
| **Yale New Haven Hospital** | 10860 | 24.9 (19.7-31.5) | 0.899 (0.889-0.909) | 0.346 | 7.6% | 90.3% (89.7-90.8) | 72.9% (72.1-73.8) | 21.4% (20.6-22.2) | 98.9% (98.7-99.1) |
| **External Validation Sites** | |  |  |  |  |  |  |  |  |
| **Bridgeport Hospital** | 17915 | 19.2 (16.2-22.8) | 0.879 (0.870-0.886) | 0.353 | 9.9% | 91.5% (91.1-91.9) | 64.2% (63.4-64.9) | 21.9% (21.3-22.5) | 98.6% (98.4-98.7) |
| **Greenwich Hospital** | 4306 | 15.6 (11.3-21.4) | 0.866 (0.846-0.884) | 0.334 | 8.5% | 87.8% (86.8-88.8) | 68.4% (67.0-69.8) | 20.6% (19.4-21.8) | 98.4% (98.0-98.7) |
| **Lawrence + Memorial Hospital** | 17730 | 21.4 (17.9-25.6) | 0.887 (0.878-0.895) | 0.338 | 8.3% | 90.7% (90.2-91.1) | 68.8% (68.1-69.5) | 20.8% (20.2-21.4) | 98.8% (98.6-99.0) |
| **Westerly Hospital** | 3614 | 13.9 (10.2-19.0) | 0.869 (0.849-0.887) | 0.371 | 10.7% | 87.8% (86.8-88.9) | 65.8% (64.3-67.4) | 23.5% (22.1-24.9) | 97.8% (97.4-98.3) |
| **ELSA-Brasil** | 3012 | 51.1 (22.2-117.7) | 0.922 | 0.201 | 1.2% | 81.1% (79.7-82.5) | 92.3% (91.3-93.2) | 11.5% (10.4-12.7) | 99.7% (99.6-99.9) |

**Abbreviations:** AUROC, area under the receiver operating characteristic curve; NPV, negative predictive value; OR, odds ratio; PPV, positive predictive value.

### **Table S12. Performance Measures of the Label-specific Convolutional Neural Network Model for Detecting Moderate or Severe Left-sided Valvular Disease in the Held-out Test Set and Across External Validation Sites**

| **Subgroup** | **Total Number** | **Diagnostic OR** | **AUROC** | **F1 Score** | **Prevalence** | **Sensitivity** | **Specificity** | **PPV** | **NPV** |
| --- | --- | --- | --- | --- | --- | --- | --- | --- | --- |
| **Held-out Test Set** |  |  |  |  |  |  |  |  |  |
| **Yale New Haven Hospital** | 7426 | 7.1 (6.0-8.5) | 0.789 (0.776-0.802) | 0.449 | 21.1% | 90.4% (89.7-91.0) | 43.1% (42.0-44.2) | 29.9% (28.8-30.9) | 94.4% (93.8-94.9) |
| **External Validation Sites** | |  |  |  |  |  |  |  |  |
| **Bridgeport Hospital** | 11243 | 7.0 (6.0-8.1) | 0.763 (0.753-0.772) | 0.503 | 27.2% | 93.2% (92.7-93.6) | 33.9% (33.0-34.7) | 34.4% (33.5-35.3) | 93.0% (92.5-93.5) |
| **Greenwich Hospital** | 3477 | 9.6 (7.1-13.1) | 0.790 (0.774-0.805) | 0.502 | 26.6% | 95.0% (94.3-95.7) | 33.5% (31.9-35.0) | 34.1% (32.5-35.6) | 94.9% (94.2-95.6) |
| **Lawrence + Memorial Hospital** | 13330 | 6.0 (5.2-6.9) | 0.760 (0.750-0.771) | 0.401 | 19.8% | 92.2% (91.7-92.6) | 33.7% (32.9-34.5) | 25.6% (24.8-26.3) | 94.6% (94.2-95.0) |
| **Westerly Hospital** | 2171 | 9.1 (6.4-13.0) | 0.790 (0.771-0.811) | 0.611 | 37.7% | 95.7% (94.9-96.6) | 28.9% (27.0-30.8) | 44.9% (42.8-47.0) | 91.8% (90.6-92.9) |
| **ELSA-Brasil** | 2987 | 10.1 (4.0-25.4) | 0.810 | 0.064 | 1.8% | 90.9% (89.9-91.9) | 50.2% (48.4-52.0) | 3.3% (2.7-4.0) | 99.7% (99.5-99.9) |

**Abbreviations:** AUROC, area under the receiver operating characteristic curve; NPV, negative predictive value; OR, odds ratio; PPV, positive predictive value.

### **Table S13. Performance Measures of the Label-specific Convolutional Neural Network Model for Detecting Moderate or Severe Aortic Regurgitation in the Held-out Test Set and Across External Validation Sites**

| **Subgroup** | **Total Number** | **Diagnostic OR** | **AUROC** | **F1 Score** | **Prevalence** | **Sensitivity** | **Specificity** | **PPV** | **NPV** |
| --- | --- | --- | --- | --- | --- | --- | --- | --- | --- |
| **Held-out Test Set** |  |  |  |  |  |  |  |  |  |
| **Yale New Haven Hospital** | 10026 | 4.0 (2.8-5.8) | 0.686 (0.659-0.712) | 0.093 | 3.9% | 91.3% (90.8-91.9) | 27.8% (26.9-28.7) | 4.9% (4.5-5.3) | 98.7% (98.5-99.0) |
| **External Validation Sites** | |  |  |  |  |  |  |  |  |
| **Bridgeport Hospital** | 15786 | 3.3 (2.5-4.3) | 0.663 (0.643-0.683) | 0.098 | 4.4% | 91.3% (90.8-91.7) | 24.1% (23.4-24.8) | 5.2% (4.8-5.5) | 98.4% (98.2-98.6) |
| **Greenwich Hospital** | 4248 | 4.2 (2.5-7.1) | 0.684 (0.649-0.716) | 0.118 | 5.3% | 93.3% (92.6-94.1) | 23.1% (21.9-24.4) | 6.3% (5.6-7.1) | 98.4% (98.0-98.8) |
| **Lawrence + Memorial Hospital** | 16663 | 2.7 (2.1-3.5) | 0.656 (0.636-0.676) | 0.093 | 4.2% | 89.8% (89.3-90.2) | 23.6% (23.0-24.3) | 4.9% (4.5-5.2) | 98.2% (98.0-98.4) |
| **Westerly Hospital** | 3505 | 4.8 (2.4-9.4) | 0.683 (0.647-0.726) | 0.118 | 5.4% | 95.2% (94.5-95.9) | 19.4% (18.1-20.8) | 6.3% (5.5-7.1) | 98.6% (98.2-99.0) |
| **ELSA-Brasil** | 3002 | - | 0.804 | 0.022 | 0.8% | 100.0% | 26.0% (24.5-27.6) | 1.1% (0.7-1.4) | 100.0% |

**Abbreviations:** AUROC, area under the receiver operating characteristic curve; NPV, negative predictive value; OR, odds ratio; PPV, positive predictive value.

### **Table S14. Performance Measures of the Label-specific Convolutional Neural Network Model for Detecting Moderate or Severe Aortic Stenosis in the Held-out Test Set and Across External Validation Sites**

| **Subgroup** | **Total Number** | **Diagnostic OR** | **AUROC** | **F1 Score** | **Prevalence** | **Sensitivity** | **Specificity** | **PPV** | **NPV** |
| --- | --- | --- | --- | --- | --- | --- | --- | --- | --- |
| **Held-out Test Set** |  |  |  |  |  |  |  |  |  |
| **Yale New Haven Hospital** | 7711 | 7.3 (5.4-9.8) | 0.793 (0.771-0.814) | 0.165 | 5.5% | 88.5% (87.8-89.2) | 48.6% (47.5-49.7) | 9.1% (8.5-9.8) | 98.6% (98.4-98.9) |
| **External Validation Sites** | |  |  |  |  |  |  |  |  |
| **Bridgeport Hospital** | 12264 | 5.8 (4.6-7.3) | 0.758 (0.740-0.774) | 0.163 | 5.8% | 87.7% (87.1-88.3) | 44.9% (44.0-45.8) | 9.0% (8.4-9.5) | 98.3% (98.1-98.6) |
| **Greenwich Hospital** | 3553 | 7.1 (4.5-11.2) | 0.771 (0.743-0.802) | 0.174 | 6.4% | 90.7% (89.8-91.7) | 42.2% (40.5-43.8) | 9.6% (8.7-10.6) | 98.5% (98.1-98.9) |
| **Lawrence + Memorial Hospital** | 13524 | 6.0 (4.7-7.8) | 0.762 (0.742-0.780) | 0.125 | 4.5% | 89.5% (88.9-90.0) | 41.6% (40.8-42.4) | 6.7% (6.3-7.1) | 98.8% (98.6-99.0) |
| **Westerly Hospital** | 2530 | 8.3 (4.9-13.8) | 0.728 (0.697-0.756) | 0.233 | 9.3% | 93.2% (92.2-94.2) | 37.5% (35.6-39.4) | 13.3% (12.0-14.6) | 98.2% (97.7-98.7) |
| **ELSA-Brasil** | 3005 | - | 0.886 | 0.006 | 0.1% | 100.0% | 50.7% (49.0-52.5) | 0.3% (0.1-0.5) | 100.0% |

**Abbreviations:** AUROC, area under the receiver operating characteristic curve; NPV, negative predictive value; OR, odds ratio; PPV, positive predictive value.

### **Table S15. Performance Measures of the Label-specific Convolutional Neural Network Model for Detecting Moderate or Severe Mitral Regurgitation in the Held-out Test Set and Across External Validation Sites**

| **Subgroup** | **Total Number** | **Diagnostic OR** | **AUROC** | **F1 Score** | **Prevalence** | **Sensitivity** | **Specificity** | **PPV** | **NPV** |
| --- | --- | --- | --- | --- | --- | --- | --- | --- | --- |
| **Held-out Test Set** |  |  |  |  |  |  |  |  |  |
| **Yale New Haven Hospital** | 10264 | 7.3 (5.7-9.3) | 0.773 (0.758-0.787) | 0.232 | 9.3% | 92.2% (91.7-92.7) | 38.3% (37.3-39.2) | 13.3% (12.7-14.0) | 97.9% (97.7-98.2) |
| **External Validation Sites** | |  |  |  |  |  |  |  |  |
| **Bridgeport Hospital** | 17000 | 6.5 (5.5-7.7) | 0.761 (0.751-0.772) | 0.28 | 12.4% | 92.8% (92.4-93.2) | 33.5% (32.8-34.2) | 16.5% (15.9-17.0) | 97.0% (96.8-97.3) |
| **Greenwich Hospital** | 4410 | 10.8 (7.3-16.1) | 0.777 (0.758-0.795) | 0.316 | 14.2% | 95.9% (95.3-96.4) | 31.9% (30.5-33.2) | 18.9% (17.7-20.1) | 97.9% (97.5-98.3) |
| **Lawrence + Memorial Hospital** | 17044 | 6.2 (5.1-7.4) | 0.755 (0.743-0.767) | 0.227 | 9.8% | 92.9% (92.5-93.3) | 32.1% (31.4-32.8) | 12.9% (12.4-13.4) | 97.6% (97.4-97.9) |
| **Westerly Hospital** | 3612 | 7.6 (5.1-11.3) | 0.774 (0.754-0.794) | 0.313 | 14.9% | 95.2% (94.5-95.9) | 27.8% (26.3-29.3) | 18.7% (17.4-20.0) | 97.0% (96.5-97.6) |
| **ELSA-Brasil** | 2999 | 7.3 (2.5-21.0) | 0.832 | 0.035 | 1.0% | 86.7% (85.5-87.9) | 52.9% (51.1-54.7) | 1.8% (1.3-2.3) | 99.7% (99.6-99.9) |

**Abbreviations:** AUROC, area under the receiver operating characteristic curve; NPV, negative predictive value; OR, odds ratio; PPV, positive predictive value.

### **Table S16. Performance Measures of the Label-specific Convolutional Neural Network Model for Detecting Severe Left Ventricular Hypertrophy in the Held-out Test Set and Across External Validation Sites**

| **Subgroup** | **Total Number** | **Diagnostic OR** | **AUROC** | **F1 Score** | **Prevalence** | **Sensitivity** | **Specificity** | **PPV** | **NPV** |
| --- | --- | --- | --- | --- | --- | --- | --- | --- | --- |
| **Held-out Test Set** |  |  |  |  |  |  |  |  |  |
| **Yale New Haven Hospital** | 6577 | 33.7 (8.1-141.0) | 0.890 (0.843-0.932) | 0.03 | 0.5% | 93.9% (93.4-94.5) | 68.5% (67.4-69.6) | 1.5% (1.2-1.8) | 100.0% (99.9-100.0) |
| **External Validation Sites** | |  |  |  |  |  |  |  |  |
| **Bridgeport Hospital** | 9408 | 9.6 (5.6-16.5) | 0.826 (0.792-0.857) | 0.052 | 1.4% | 88.7% (88.1-89.4) | 55.0% (54.0-56.0) | 2.7% (2.4-3.1) | 99.7% (99.6-99.8) |
| **Greenwich Hospital** | 1741 | 12.0 (2.7-53.5) | 0.846 (0.708-0.957) | 0.041 | 0.9% | 86.7% (85.1-88.3) | 64.9% (62.7-67.2) | 2.1% (1.4-2.8) | 99.8% (99.6-100.0) |
| **Lawrence + Memorial Hospital** | 11876 | 44.2 (10.8-181.0) | 0.899 (0.864-0.929) | 0.024 | 0.5% | 96.7% (96.3-97.0) | 60.4% (59.5-61.3) | 1.2% (1.0-1.4) | 100.0% (99.9-100.0) |
| **Westerly Hospital** | 1630 | 12.9 (3.0-55.9) | 0.846 (0.761-0.913) | 0.052 | 1.2% | 90.0% (88.5-91.5) | 58.9% (56.6-61.3) | 2.7% (1.9-3.4) | 99.8% (99.6-100.0) |
| **ELSA-Brasil** | 3014 | 5.7 (1.0-31.1) | 0.773 | 0.01 | 0.2% | 66.7% (65.0-68.3) | 74.0% (72.4-75.5) | 0.5% (0.3-0.8) | 99.9% (99.8-100.0) |

**Abbreviations:** AUROC, area under the receiver operating characteristic curve; NPV, negative predictive value; OR, odds ratio; PPV, positive predictive value.

### **Table S17. Baseline Characteristics of Individuals Eligible for the Predictive Assessment of ADAPT-HEART in Yale New Haven Hospital, Hospital-based External Validation Sites, and the UK Biobank**

| **Characteristic** | **Yale New Haven Hospital** | **Bridgeport Hospital** | **Greenwich Hospital** | **Lawrence + Memorial Hospital** | **Westerly Hospital** | **UK Biobank** |
| --- | --- | --- | --- | --- | --- | --- |
| **Number** | 127,547 | 46,883 | 26,835 | 28,344 | 3,930 | 41,800 |
| **Age (years)** | 53 [37-67] | 53 [38-67] | 59 [45-74] | 59 [43-72] | 63 [51-74] | 65 [59-71] |
| **Sex** | 73031 (57.3%) | 27588 (58.8%) | 15443 (57.5%) | 16314 (57.6%) | 2138 (54.4%) | 21671 (51.8%) |
| **Race/Ethnicity** |  |  |  |  |  |  |
| White | 75450 (60.7%) | 19597 (42.8%) | 19002 (72.8%) | 20609 (74.4%) | 3596 (92.8%) | 40359 (96.8%) |
| Black | 24481 (19.7%) | 11519 (25.1%) | 1356 (5.2%) | 2448 (8.8%) | 71 (1.8%) | 300 (0.7%) |
| Asian | 2591 (2.1%) | 493 (1.1%) | 726 (2.8%) | 392 (1.4%) | 31 (0.8%) | 598 (1.4%) |
| Hispanic | 20217 (16.3%) | 13814 (30.1%) | 4848 (18.6%) | 3724 (13.4%) | 127 (3.3%) |  |
| Others | 1471 (1.2%) | 412 (0.9%) | 174 (0.7%) | 516 (1.9%) | 50 (1.3%) | 430 (1.0%) |
| **Hypertension** | 56313 (44.2%) | 21259 (45.3%) | 10705 (39.9%) | 14437 (50.9%) | 2289 (58.2%) | 5941 (14.2%) |
| **DM** | 21355 (16.7%) | 9776 (20.9%) | 3830 (14.3%) | 5485 (19.4%) | 855 (21.8%) | 1224 (2.9%) |
| **Incident SHD** | 5353 (4.2%) | 3507 (7.5%) | 1493 (5.6%) | 2290 (8.1%) | 298 (7.6%) | 413 (1.0%) |
| **TTE-defined SHD** | 4178 (3.3%) | 2880 (6.1%) | 1021 (3.8%) | 1810 (6.4%) | 221 (5.6%) |  |
| **HF Hospitalization** | 1751 (1.4%) | 1229 (2.6%) | 761 (2.8%) | 876 (3.1%) | 138 (3.5%) | 44 (0.1%) |
| **Aortic Valve Disease** | 518 (0.4%) | 172 (0.4%) | 48 (0.2%) | 72 (0.3%) | 10 (0.3%) | 228 (0.5%) |
| **Mitral Valve Disease** | 199 (0.2%) | 55 (0.1%) | 20 (0.1%) | 27 (0.1%) | 4 (0.1%) | 264 (0.6%) |
| **Follow-up (years)** | 4.0 [1.7-6.4] | 4.2 [2.4-6.2] | 4.7 [2.7-6.5] | 2.5 [1.1-4.1] | 2.4 [0.8-4.0] | 3.0 [2.1-4.5] |

**Abbreviations:** DM, diabetes mellitus; SHD, structural heart disease; TTE, transthoracic echocardiogram.

### **Figure S1. The Study Flow Diagram**

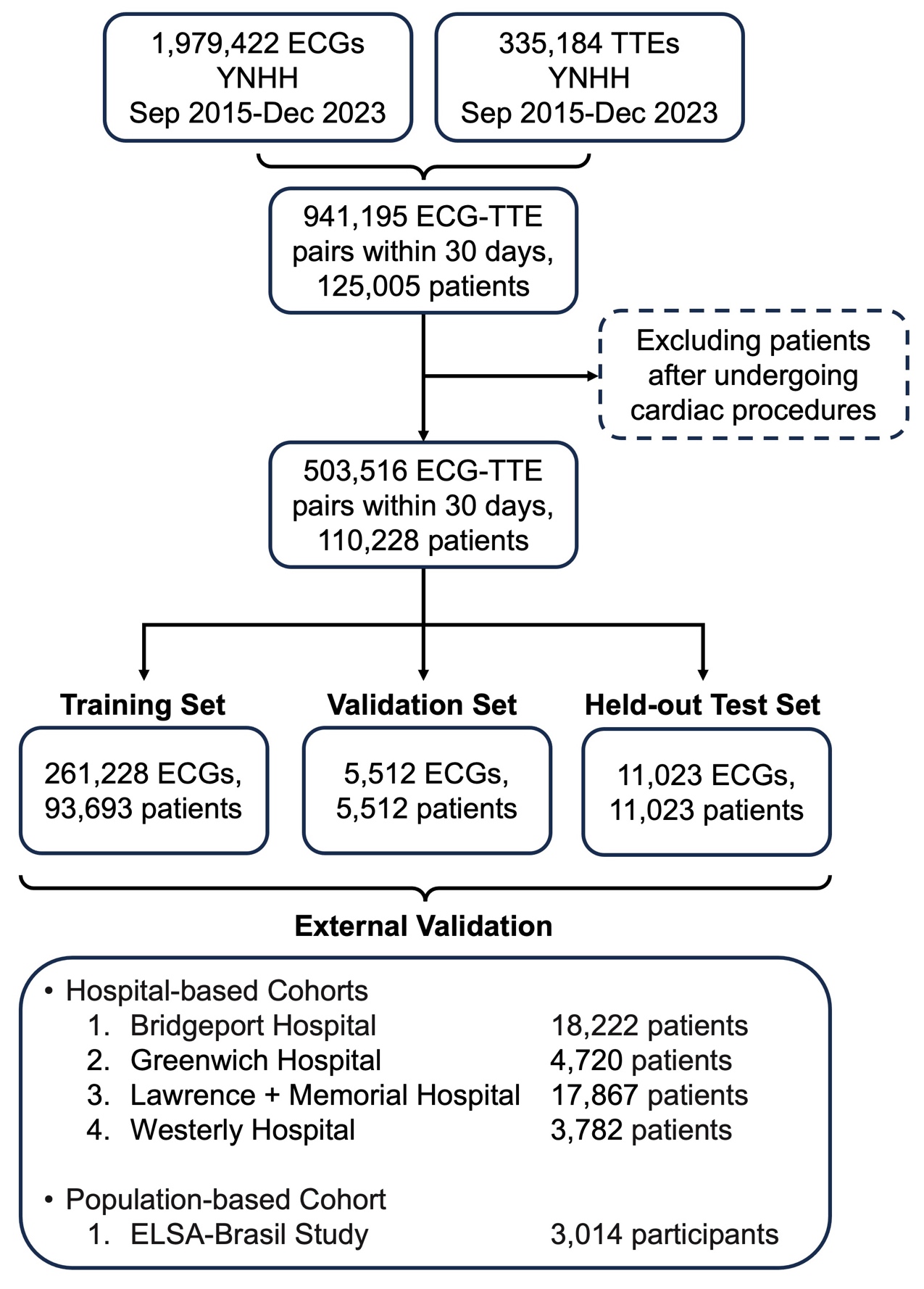

**Abbreviations:** ECG, electrocardiogram; TTE, transthoracic echocardiogram; YNHHS, Yale New Haven Health System.

### **Figure S2. Receiver Operating Characteristic Curves of the ADAPT-HEART for Detecting Individual Structural Heart Disease in the Internal Held-out Test Set and External Validation Cohorts**

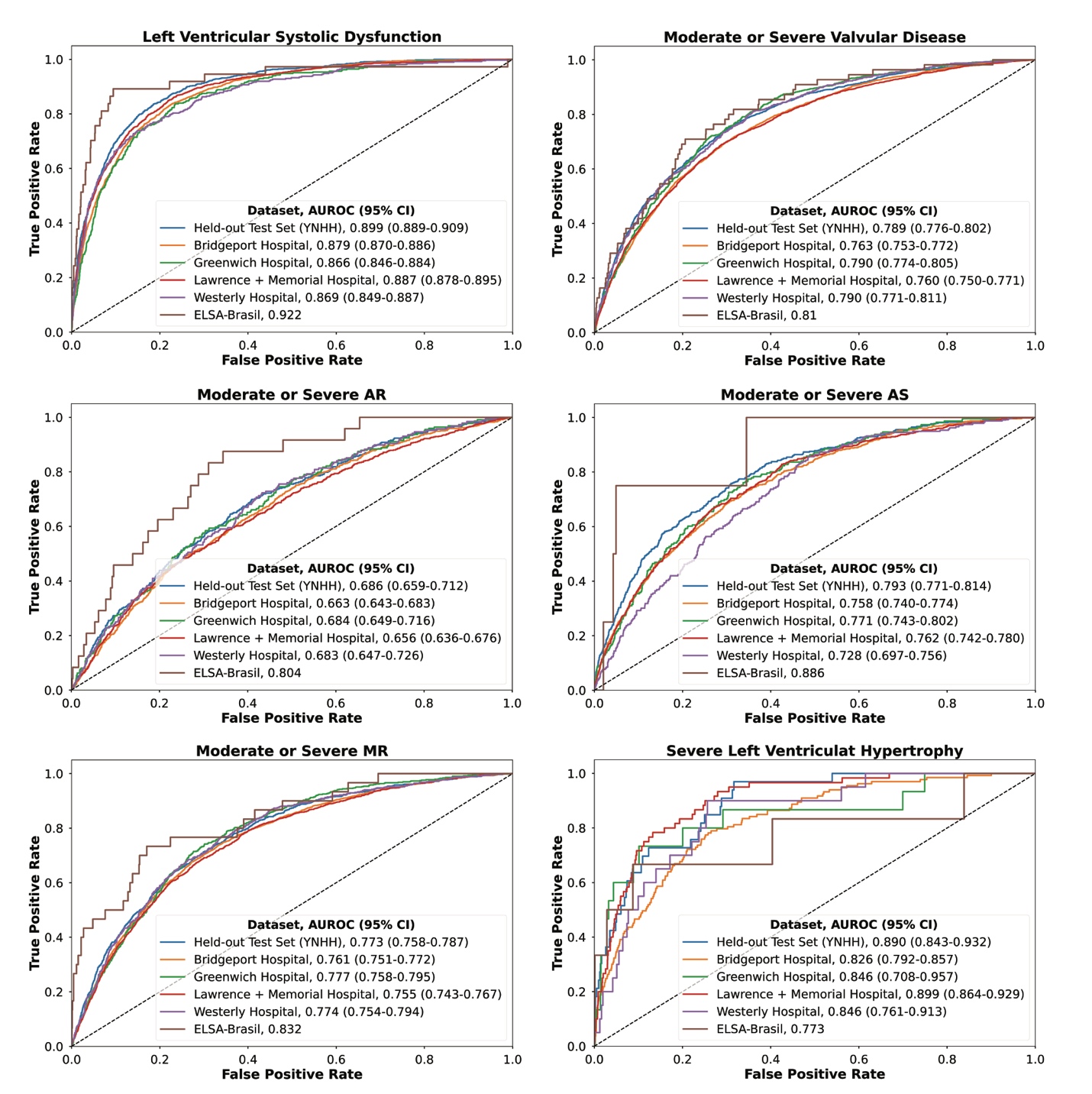

**Abbreviations:** ADAPT-HEART, AI Deep learning for Adapting Portable Technology in HEART disease detection; AR, aortic regurgitation; AS, aortic stenosis; AUROC, area under the receiver operating characteristic curve; LVEF, left ventricular ejection fraction; MR, mitral regurgitation; YNHH, Yale New Haven Hospital.
